# Incidence and prevalence of obstructive sleep apnoea and narcolepsy in the UK: a population-based descriptive study

**DOI:** 10.1101/2025.06.18.25329848

**Authors:** Helen Strongman, Martina Sykorova, Yi Ting Yu, Aurelien Belot, Hema Mistry, Ellen Nolte, Sofia H Eriksson, Michelle A Miller, Krishnan Bhaskaran, Ian E Smith, Charlotte Warren-Gash

## Abstract

Obstructive sleep apnoea and narcolepsy affect approximately 4.8% and 0.047% of the UK population, respectively. We do not know how many people have been diagnosed or how this varies over time and by demographic factors. We therefore conducted a historical population-based descriptive study estimating prevalence and incidence of diagnosed OSA and narcolepsy in England from 2000 to 2019 stratified by demographic factors, and compared estimates to Scotland, Wales and Northern Ireland. Data were from Clinical Practice Research Datalink (CPRD) primary care records linked to Hospital Episode Statistics (HES) admissions. The study population included people with ≥ 90 days follow-up between 01/10/2000-31/12/2019, no prior record of primary or central sleep apnoea, and aged ≥18 years (OSA only). Diagnoses were defined using the first coded record for each condition in CPRD or HES data. Annual prevalence was estimated at mid-year and directly age/sex-standardised to the national population. Incidence was estimated by dividing new diagnoses by total person-time at risk. In England, 2019 adult standardised diagnosed OSA prevalence was 1.40% (95% CI 1.40-1.41) representing approximately 622,528 people; standardised narcolepsy prevalence was 0.020% (0.019-0.021) representing approximately 11,307 people. Despite increases over time, diagnosed incidence and prevalence remained lower than published estimates of symptomatic frequency. Rates varied by age, sex, ethnicity and UK nation for both conditions, and urban-rural living, area-based deprivation and practice size for OSA. Our results call for high quality research to drive initiatives that increase diagnosis rates and address variation.

**Graphical Abstract:** 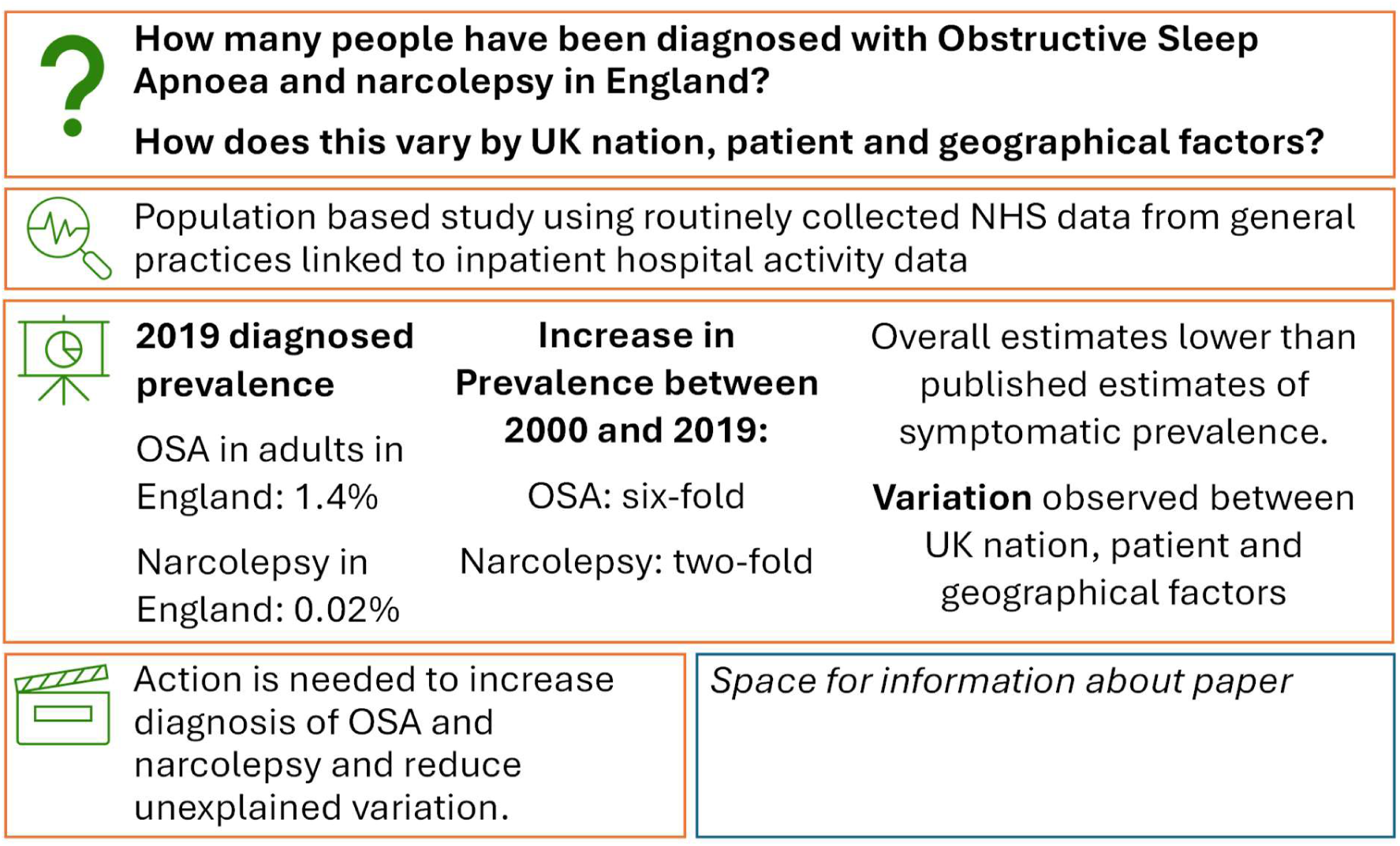

## INTRODUCTION

Obstructive sleep apnoea (OSA) and narcolepsy are well-characterised sleep disorders that affect people’s quality of life, physical and psychological health (1). OSA is a respiratory condition associated with oxygen desaturation at night, snoring and gasping, and is sometimes referred to as obstructive sleep apnoea/hypopnoea syndrome (OSAHS). Narcolepsy is a neurological condition affecting the brain’s ability to regulate sleep and wakefulness. Both conditions may present with excessive daytime sleepiness and disrupted nighttime sleep (1). They are diagnosed in specialist care, often in sleep centres led by respiratory specialists or neurologists. The advent of improved diagnostic devices may enable diagnosis in primary care in future(2).

Systematic reviews have revealed widespread variation in international estimates of narcolepsy and OSA frequency due to methodological variation and limitations of individual studies. Estimates range from 0.06 to 6.56 per 100,000 person-years for narcolepsy incidence (3), 1.05 to 79.40 per 100,000 population for narcolepsy prevalence(3), 9% to 38% and 6% to 17% for mild-to-severe and moderate-to-severe symptomatic OSA prevalence, respectively(4).

In the United Kingdom (UK), the symptomatic prevalence of moderate to severe OSA is estimated to be 4.8%, based on predictive methods using estimates from other countries (5); the symptomatic prevalence of narcolepsy is estimated to be 0.047% (95% CI 0.016-0.078) based on questionnaire studies from the 1990s (6,7). Routinely collected healthcare records collected from primary and secondary care providers in the UK have recently been shown to ascertain incident and prevalent OSA cases with high accuracy, and prevalent narcolepsy cases with moderate accuracy although delayed recording of narcolepsy diagnoses limits our ability to ascertain incident narcolepsy cases(8). These data have previously been used to estimate incidence of diagnosed narcolepsy(9–11) and prevalence of diagnosed OSA(12). To our knowledge, however, there are no UK studies estimating prevalence of diagnosed narcolepsy, incidence of diagnosed OSA, or describing variation in the diagnosed frequency of these conditions within the UK population(13).

National guidelines and commissioning decisions therefore rely on minimal epidemiological evidence regarding the diagnosed prevalence and incidence of these conditions and how this varies by key risk factors(14,15). More detailed knowledge of diagnosed prevalence and incidence rates of both sleep disorders in the UK would facilitate effective OSA and narcolepsy management, and better allocation of healthcare resources(16). We used electronic health records data to describe diagnosed prevalence and incidence of OSA and narcolepsy in England, examine variation over time and by demographic characteristics, and compare overall estimates to the devolved nations of the UK. We assessed the robustness of this evidence based on our recently published validation study(8).

## METHODS

### Study design and setting

In this population-based descriptive study, we used routinely collected data provided by Clinical Practice Research Datalink (CPRD). We combined primary care electronic health record data from CPRD Aurum and CPRD GOLD(17), which include people registered in National Health Service (NHS) practices that use Vision and EMIS software systems, respectively. Primary care staff use these software systems to record data for clinical and administrative purposes. Clinical events, including diagnoses, are coded using Read codes (Vision/GOLD) or Systematised Nomenclature of Medicine Clinical Terms (Snomed-CT codes - (EMIS/Aurum).

Data for most English practices and patients are linked to multiple data sets(18). We used ICD-10 coded Hospital Episodes Statistics (HES) Admitted Patient Care (APC) data(19), Office of National Statistics (ONS) mortality death dates (20), practice level small area-based deprivation data, and patient level small-area based urban rural data (21). HES APC is a national data collection on hospital activity used to inform management, planning and payment for services. DOIs with links describing the individual datasets and data governance arrangements are provided (Supplementary Appendix table 1). Together, the versions of the two databases used for this study cover approximately 25% of the UK population.

This study was approved by the London School of Hygiene & Tropical Medicine Ethics Committee (Ref 101296) and CPRD’s Research Data Governance process (protocol 22_001887). The study protocol is available online(22). CPRD supplies anonymised data for public health research; therefore, individual patient consent was not required for this study.

### Study population

We selected people registered in contributing practices, excluding records that failed basic CPRD quality checks, did not have a recorded sex, were not eligible for linkage or were not alive and registered in the practice during the time period covered by the linked data (02/01/1998 to 29/03/2021). As data quality in CPRD data may be unreliable prior to 2000(23) and recording of medical conditions in primary care data changed during the COVID-19 pandemic(24), we analysed data from 01/01/2000 to 31 December 2019. Data from practices contributing to both CPRD GOLD and CPRD Aurum, and CPRD Aurum practices that have merged into other contributing practices, were excluded from CPRD GOLD to avoid duplication.

Existing conditions may be recorded in the primary care record shortly after a patient joins a new practice and misclassified as incident rather than prevalent cases. To avoid misclassification, we identified a minimum registration period of 90 days by visualising incidence rates of OSA and narcolepsy in the year following registration and further excluded people who left the practice or died before this minimum period(25). (Supplementary Appendix Figure 1).

### Procedures

We used validated codelists and algorithms combining primary care and HES APC data to identify OSA and narcolepsy cases(8). Codes for sleep apnoea are categorised as OSA, Obstructive sleep apnoea syndrome (OSAS), Sleep apnoea NOS, Sleep apnoea syndrome NOS, Central sleep apnoea only and Primary sleep apnoea only. Codes for mixed sleep apnoea are included in the OSA category. Variation in coding of OSA reflects differences in terminology used to describe the condition over time and in different settings(14). We defined narcolepsy diagnosis dates using the event date of the first ever coded record of narcolepsy in primary care or HES APC data; people with one or more missing event dates for a narcolepsy code were excluded from the analysis. OSA diagnoses dates were defined in the same way including all sleep apnoea categories except central and primary sleep apnoea which are distinct from OSA.

Procedures used to define prevalent, incident and unexposed follow-up time are described in Figure 1. The follow-up start date was defined as the latest of the start of the linkage coverage period, practice registration date plus 90 days, or the individual’s 18th birthday (OSA only). People with a record of primary or central sleep apnoea before the follow-up start date or aged <18 years at the start of follow-up were excluded from the OSA analyses. People were counted as prevalent cases from the latest of the follow-up start date or the sleep disorder diagnosis date; those diagnosed after the follow-up start date were counted as unexposed until the diagnosis date and both incident and prevalent cases from diagnosis. Follow-up was censored at the earliest of the end of the linkage coverage period, end of registration in practice, ONS death date, or first record of central or primary sleep apnoea (OSA only). Individuals transferring between practices may be represented in the data more than once, but at different time points.

**Figure 1:**
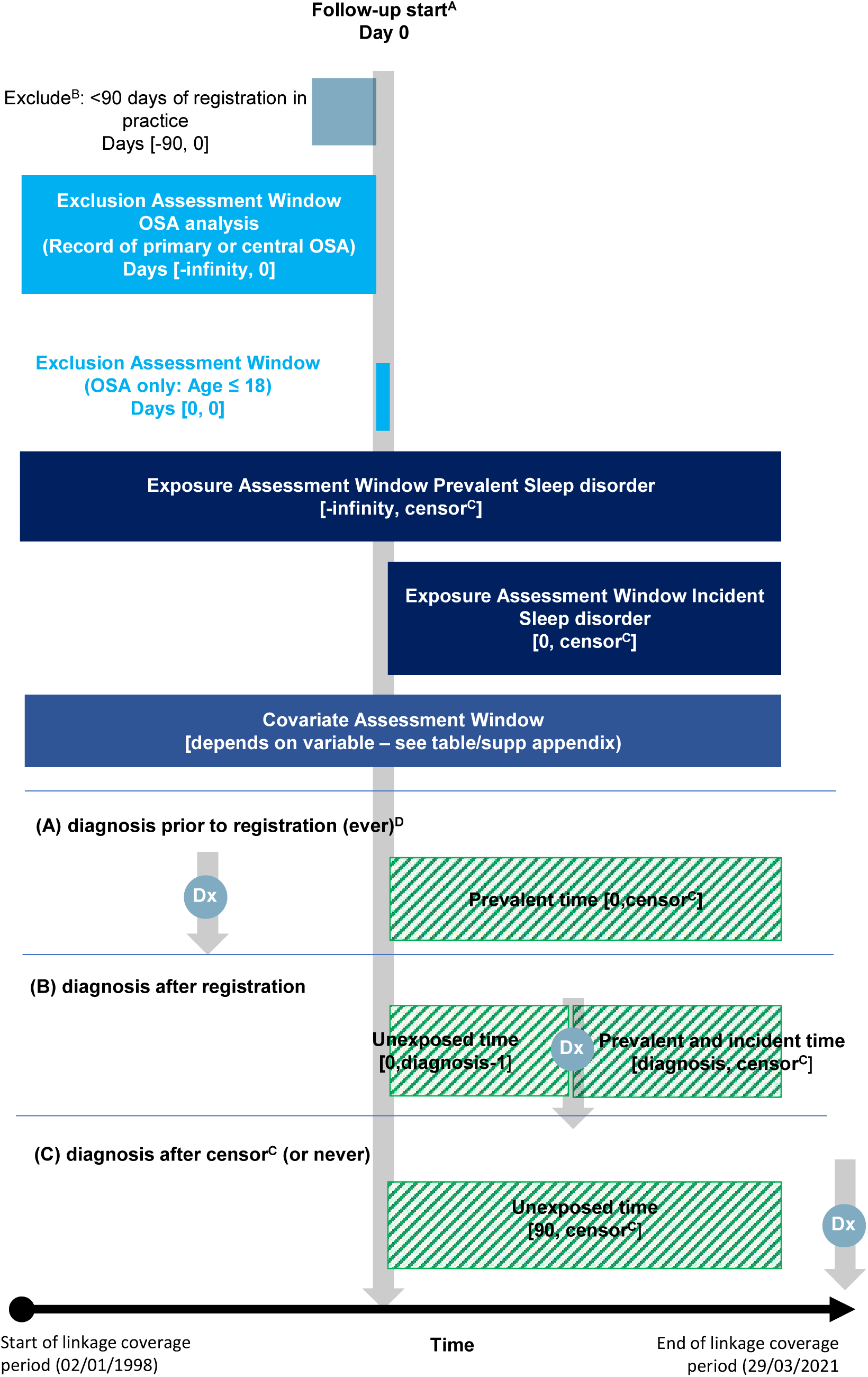
Procedures used to identify the study population and incident, prevalent and unexposed follow-up time. ^A^Follow-up start (latest of practice registration date + 90 days, start of linkage coverage period, 18th birthday – OSA only) ^B^Additional data quality exclusions (not research quality, gender not recorded, duplicate practice, not eligible for linkages – linked cohort only, >= 1 coded record for sleep disorder with missing date) ^C^censor: Earliest of end of linkage coverage period, end of registration in practice, death or record of central of primary sleep apnoea (OSA only) ^D^Excluded from study population for incidence analysis (not at risk) OSA = Obstructive Sleep Apnoea Created from open access template (Schneeweiss S, Rassen JA, Brown JS, Rothman KJ, Happe L, Arlett P, Dal Pan G, Goettsch W, Murk W, Wang SV. Graphical Depiction of Longitudinal Study Designs in Health Care Databases. Ann Intern Med. 2019 Mar 19;170(6):398-406. doi: 10.7326/M18-3079. Epub 2019 Mar 12. PMID: 30856654.)

Fixed covariates were identified from the following data sources: sex (practice registration data at the time of data collection), ethnicity (most common ethnicity recorded in primary care or latest if equally common, unknown or missing values replaced with most commonly available record in linked HES data where available), area-based deprivation quintile (Carstairs index from 2011 census mapped to practice postcode), urban-rural (urban-rural category from 2011 census mapped to patient postcode at time of linkage), region (ONS region for practices within England), practice size (estimated from CPRD denominator population registered in the practice on 01/07/2019). Age at the mid-point of each year was imputed from practice registration data (01/07/year of birth) and grouped as 9 to <18 (narcolepsy only), 18 to < 25, 10-year categories from 25 to <85 and greater than 85. In the OSA study population, BMI (OSA only) at the mid-point of each year was estimated using the latest available record and grouped into 6-level World Health Organisation (WHO) BMI categories (underweight to Obesity class III +) Data management and statistical analysis were done in Stata MP, version 18. Full data management and programming code can be found at [INSERT GITHUB LINK WHEN AVAILABLE]. Updated codelists are available at [INSERT LSHTM DATACOMPASS LINK WHEN AVAILABLE]. Full sleep disorder and covariate definitions are available in Strongman et al 2025(8).

### Statistical analysis

We estimated crude annual mid-year point prevalence and 95% confidence intervals (CIs, via exact binomial methods) for each sleep disorder from 01/01/2000 to 31/12/2019. We directly standardised age and sex specific prevalence to ONS population figures(26) for each year, estimating 95% standardised CIs using Poisson processes.. The number of people with prevalent diagnoses each year and related plausibility intervals were estimated by multiplying the ONS population for each year to the standardised rates and CI bounds.

Crude incidence rates were calculated for each year for OSA and for 5-calendar year categories for narcolepsy to avoid small numbers; 95% CIs were estimated assuming a Poisson distribution for the number of cases.

To describe diagnosed frequency by demographic characteristics, we estimated prevalence rates (for 2019) and incidence rates (for each five-year category, and 2019 for OSA) across all covariate values. To examine variation in diagnosed frequency by each covariate, we estimated prevalence ratios using log binomial models(27) and incidence rate ratios using Poisson models. All models were adjusted by 5-year age groups and sex for both sleep disorders, and for categorical BMI for OSA only.

### Secondary analyses

We repeated the study using primary care data only to compare diagnosed incidence and prevalence of OSA and narcolepsy in Scotland, Wales and Northern Ireland with England. In defining our study population, we used the CPRD derived death date in place of the ONS death date. OSA and narcolepsy cases were identified using primary care data only.

### Sensitivity analyses

Our previous work demonstrated that case definitions using only primary care data identified fewer true and false cases than those incorporating both primary care and linked hospital data(8). We therefore estimated incidence and prevalence rates over time and prevalence and incidence rate ratios in England for the primary care only definition as a sensitivity analysis.

People who were underweight were excluded from analyses estimating incidence rates, incidence rate ratios and prevalence ratios due to small numbers. All models used complete case data; to investigate the impact of this on BMI-adjusted models we ran age-sex adjusted models restricted to people with complete BMI data.

## RESULTS

### Full study population

The initial study population included 37,501,230 people who were alive and registered in CPRD practices for at least 90 days and during the linkage coverage period, met basic data quality checks and were eligible for linkage (Supplementary Appendix Figure 2). Of these, 475,850 and 8,947 had coded records for OSA and narcolepsy, respectively. After excluding people with diagnosis dates after the end of the study or follow-up period, missing dates for at least one sleep disorder record, and people with primary or central sleep apnoea before the start of follow-up or aged <18 years at the end of follow-up for the OSA study population, 270,100 and 6,914 had prevalent OSA or narcolepsy during the linkage coverage period, respectively, 191,220 and 2,919 of whom were diagnosed during follow-up (i.e. incident cases).

### Who had diagnosed OSA and narcolepsy in 2019 and who was diagnosed in this period?

The estimated OSA crude incidence rate, standardised prevalence and number of cases of OSA in the 2019 adult population of England were 170.9 per 100 000 person-years (95% CI 168.4-173.5), 1.406% (95% CI 1.399-1.414), and 622,528 (95% plausible interval 619,336-625,720) respectively. The estimated crude incidence rate of narcolepsy in England from 2015-2019 was 1.178 per 100,000 person-years (95% CI 1.097-1.265); the 2019 standardised prevalence was 0.020 (0.019-0.021) equating to 11,307 people with narcolepsy in the population (95% plausible interval 10,876-11,738).

Table 1 describes the composition of the OSA and narcolepsy groups in 2019 (2014-2019 for narcolepsy incidence rates) by patient and area-based characteristics, and crude prevalence and incidence rates for each characteristic. Overall crude prevalence was similar to standardised prevalence as described above. The composition of the denominator populations, including numbers of practices in each region, is described in Supplementary Appendix table 2. As multiple regions were represented by small numbers of practices, we have not analysed variation by region further. Median age for adults with prevalent OSA and narcolepsy, respectively, was 59 (Inter-quartile range (IQR) 49,68) and 47 years (IQR 34,63); 29.5% (n=42,228) and 53.2% (1417) were female. The estimated median age at diagnosis was 54 (IQR 44-65) and 34 years (IQR 23-49). While the majority of people with prevalent OSA were classified as obese, 0.3% were underweight (n=432), 8.4% were of normal weight (n=11,986) and 23.7% (n=33,904) were overweight. Incidence rates and prevalence of OSA peaked in people aged 55-<65 and 65-75 years, respectively. Incidence rates for narcolepsy peaked in the 18-<25 year age groups decreasing thereafter, whereas prevalence increased steadily until the 35-45 year age group and then plateaued; there was weak statistical evidence that both incidence and prevalence increased slightly in the oldest age groups.

**Table 1:** Prevalence and incidence rates of sleep disorders in England by patient and area-based characteristics, A: Obstructive Sleep Apnoea in adults (OSA) (2019), B: Narcolepsy in the full population (2019 prevalence, 2014 to 2019 incidence) Yorks/Humber = Yorks & The Humber; BMI = Body Mass Index. Proportion of missing data in the total study population/total person years: ethnicity OSA 8.0%/8.1%, ethnicity narcolepsy 7.4%/7.1%, BMI OSA 13.6%/14.6%, Urban-rural OSA and narcolepsy 0.1%/0.1%. *The incidence rate was not estimated for the underweight category due to small numbers of cases.

| **A (OSA)** | **Prevalent cases**<br>**n (column %)** | **Crude prevalence %**<br>**(95% CI)** | **Incident cases** | **Crude incidence rate**<br>**/100 000 person-years**<br>**(95% CI)** |
| --- | --- | --- | --- | --- |
| **Full study population** | 143277 (100.0%) | 1.383 (1.376-1.390) | 17515 (100.0%) | 170.945 (168.432-173.496) |
| **Age group** |  |  |  |  |
| 18 to < 25 | 694 (0.5%) | 0.068 (0.063-0.073) | 328 (1.9%) | 30.536 (27.404-34.026) |
| 25 to < 35 | 5737 (4.0%) | 0.296 (0.289-0.304) | 1470 (8.4%) | 76.443 (72.633-80.452) |
| 35 to < 45 | 15688 (10.9%) | 0.861 (0.848-0.875) | 2756 (15.7%) | 153.061 (147.451-158.883) |
| 45 to < 55 | 32549 (22.7%) | 1.808 (1.789-1.828) | 4225 (24.1%) | 239.505 (232.391-246.836) |
| 55 to < 65 | 40265 (28.1%) | 2.601 (2.576-2.626) | 4331 (24.7%) | 287.747 (279.303-296.445) |
| 65 to < 75 | 31837 (22.2%) | 2.693 (2.664-2.723) | 2880 (16.4%) | 250.820 (241.825-260.150) |
| 75 to < 85 | 13910 (9.7%) | 1.894 (1.863-1.925) | 1291 (7.4%) | 179.432 (169.907-189.492) |
| 85 plus | 2597 (1.8%) | 0.826 (0.795-0.859) | 234 (1.3%) | 75.167 (66.128-85.442) |
| Median age (IQR) | 59 (49-68) |  | 54 (44-65) |  |
| **Sex** |  |  |  |  |
| male | 101049 (70.5%) | 1.952 (1.940-1.964) | 11232 (64.1%) | 220.554 (216.513-224.671) |
| female | 42228 (29.5%) | 0.814 (0.807-0.822) | 6283 (35.9%) | 121.921 (118.943-124.973) |
| **Ethnicity** |  |  |  |  |
| 0. White | 123610 (86.3%) | 1.559 (1.550-1.567) | 14786 (84.4%) | 189.016 (185.994-192.088) |
| 1. South Asian | 8162 (5.7%) | 1.117 (1.093-1.142) | 1166 (6.7%) | 160.557 (151.601-170.042) |
| 2. Black | 5753 (4.0%) | 1.364 (1.329-1.399) | 703 (4.0%) | 167.668 (155.721-180.532) |
| 3. Other | 2158 (1.5%) | 0.708 (0.679-0.739) | 308 (1.8%) | 101.582 (90.848-113.584) |
| 4. Mixed | 1440 (1.0%) | 1.011 (0.959-1.064) | 192 (1.1%) | 135.001 (117.194-155.513) |
| Missing | 2154 (1.5%) |  | 360 (2.1%) |  |
| **Area based deprivation** |  |  |  |  |
| 1 (least deprived) | 18673 (13.0%) | 1.437 (1.416-1.457) | 2144 (12.2%) | 166.453 (159.555-173.650) |
| 2 | 27961 (19.5%) | 1.353 (1.337-1.368) | 3372 (19.3%) | 164.089 (158.643-169.722) |
| 3 | 32279 (22.5%) | 1.374 (1.360-1.389) | 3851 (22.0%) | 166.323 (161.152-171.660) |
| 4 | 31323 (21.9%) | 1.446 (1.430-1.462) | 3884 (22.2%) | 182.159 (176.519-187.978) |
| 5 (most deprived) | 33041 (23.1%) | 1.333 (1.319-1.348) | 4264 (24.3%) | 173.662 (168.527-178.953) |
| **Urban-rural** |  |  |  |  |
| urban | 119129 (83.1%) | 1.357 (1.349-1.365) | 14908 (85.1%) | 171.577 (168.844-174.353) |
| rural | 24034 (16.8%) | 1.528 (1.509-1.548) | 2593 (14.8%) | 167.566 (161.239-174.141) |
| Missing | 114 (0.1%) |  | 14 (0.1%) |  |
| **Practice size quintiles** |  |  |  |  |
| 1 largest | 50437 (35.2%) | 1.280 (1.269-1.291) | 6225 (35.5%) | 159.654 (155.736-163.669) |
| 2 | 35019 (24.4%) | 1.452 (1.437-1.467) | 4234 (24.2%) | 177.731 (172.457-183.166) |
| 3 | 26774 (18.7%) | 1.440 (1.423-1.457) | 3242 (18.5%) | 176.084 (170.126-182.251) |
| 4 | 19693 (13.7%) | 1.468 (1.447-1.488) | 2462 (14.1%) | 186.360 (179.142-193.868) |
| 5 smallest | 11354 (7.9%) | 1.407 (1.382-1.433) | 1352 (7.7%) | 168.501 (159.755-177.727) |
| **WHO BMI category** |  |  |  |  |
| Underweight* | 432 (0.3%) | 0.183 (0.166-0.201) |  |  |
| Normal weight | 11986 (8.4%) | 0.342 (0.336-0.348) | 1682 (9.6%) | 48.119 (45.873-50.474) |
| Overweight | 33904 (23.7%) | 1.132 (1.120-1.144) | 3959 (22.6%) | 134.709 (130.578-138.972) |
| Obesity class I | 38244 (26.7%) | 2.729 (2.702-2.756) | 4275 (24.4%) | 317.810 (308.424-327.481) |
| Obesity class II | 26695 (18.6%) | 5.124 (5.064-5.184) | 3172 (18.1%) | 654.889 (632.490-678.080) |
| Obesity class III+ | 29091 (20.3%) | 10.062 (9.953-10.172) | 3645 (20.8%) | 1,448.527 (1,402.257-1,496.323) |
| Missing | 2925 (2.0%) |  | 782 (4.5%) |  |

| **B (Narcolepsy)** | **Prevalent cases**<br>**n (column %)** | **Crude prevalence %**<br>**(95% CI)** | **Incident cases** | **Crude incidence rate**<br>**/100 000 person-years**<br>**(95% CI)** |
| --- | --- | --- | --- | --- |
| **Full study population** | 2662 (100.0%) | 0.020 (0.019-0.021) | 761 (100.0%) | 1.178 (1.097-1.265) |
| **Age group** |  |  |  |  |
| 0 to < 9 | 21 (0.8%) | 0.002 (0.001-0.003) | 30 (3.9%) | 0.463 (0.324-0.663) |
| 9 to < 18 | 108 (4.1%) | 0.008 (0.006-0.009) | 88 (11.6%) | 1.339 (1.086-1.650) |
| 18 to < 25 | 168 (6.3%) | 0.014 (0.012-0.017) | 94 (12.4%) | 1.674 (1.368-2.049) |
| 25 to < 35 | 399 (15.0%) | 0.021 (0.019-0.023) | 146 (19.2%) | 1.551 (1.319-1.824) |
| 35 to < 45 | 509 (19.1%) | 0.028 (0.026-0.030) | 133 (17.5%) | 1.493 (1.260-1.769) |
| 45 to < 55 | 488 (18.3%) | 0.027 (0.025-0.030) | 106 (13.9%) | 1.158 (0.957-1.401) |
| 55 to < 65 | 364 (13.7%) | 0.024 (0.021-0.026) | 69 (9.1%) | 0.931 (0.735-1.178) |
| 65 to < 75 | 277 (10.4%) | 0.023 (0.021-0.026) | 43 (5.7%) | 0.726 (0.538-0.979) |
| 75 to < 85 | 220 (8.3%) | 0.030 (0.026-0.034) | 33 (4.3%) | 0.926 (0.659-1.303) |
| 85 plus | 108 (4.1%) | 0.034 (0.028-0.041) | 19 (2.5%) | 1.218 (0.777-1.910) |
| Median age (IQR) | 47 (34-63) |  | 34 (23-49) |  |
| **Sex** |  |  |  |  |
| male | 1245 (46.8%) | 0.019 (0.018-0.020) | 337 (44.3%) | 1.039 (0.934-1.156) |
| female | 1417 (53.2%) | 0.022 (0.021-0.023) | 424 (55.7%) | 1.318 (1.198-1.450) |
| **Ethnicity** |  |  |  |  |
| 0. White | 2261 (84.9%) | 0.023 (0.022-0.024) | 621 (81.6%) | 1.254 (1.159-1.357) |
| 1. South Asian | 77 (2.9%) | 0.008 (0.006-0.010) | 31 (4.1%) | 0.650 (0.457-0.924) |
| 2. Black | 186 (7.0%) | 0.032 (0.027-0.037) | 62 (8.1%) | 2.200 (1.715-2.821) |
| 3. Other | 28 (1.1%) | 0.007 (0.005-0.010) | 14 (1.8%) | 0.807 (0.478-1.363) |
| 4. Mixed | 53 (2.0%) | 0.021 (0.016-0.027) | 21 (2.8%) | 1.806 (1.178-2.770) |
| Missing | 57 (2.1%) |  | 12 (1.6%) |  |
| **Area based deprivation** |  |  |  |  |
| 1 (least deprived) | 325 (12.2%) | 0.020 (0.018-0.022) | 91 (12.0%) | 1.130 (0.921-1.388) |
| 2 | 507 (19.0%) | 0.020 (0.018-0.021) | 119 (15.6%) | 0.930 (0.777-1.113) |
| 3 | 613 (23.0%) | 0.021 (0.019-0.023) | 184 (24.2%) | 1.266 (1.096-1.463) |
| 4 | 604 (22.7%) | 0.022 (0.020-0.024) | 183 (24.0%) | 1.356 (1.173-1.567) |
| 5 (most deprived) | 613 (23.0%) | 0.019 (0.018-0.021) | 184 (24.2%) | 1.170 (1.013-1.352) |
| **Urban-rural** |  |  |  |  |
| urban | 2279 (85.6%) | 0.020 (0.020-0.021) | 667 (87.6%) | 1.219 (1.130-1.315) |
| Rural | 381 (14.3%) | 0.019 (0.018-0.022) | 94 (12.4%) | 0.956 (0.781-1.170) |
| Missing | 2 (0.1%) |  | 0 (0.0%) |  |
| **Practice size quintile** |  |  |  |  |
| 1 largest | 989 (37.2%) | 0.020 (0.019-0.021) | 280 (36.8%) | 1.172 (1.042-1.317) |
| 2 | 669 (25.1%) | 0.022 (0.020-0.023) | 188 (24.7%) | 1.235 (1.071-1.425) |
| 3 | 474 (17.8%) | 0.020 (0.018-0.022) | 132 (17.3%) | 1.115 (0.940-1.323) |
| 4 | 332 (12.5%) | 0.019 (0.017-0.021) | 106 (13.9%) | 1.240 (1.025-1.500) |
| 5 smallest | 198 (7.4%) | 0.019 (0.016-0.022) | 55 (7.2%) | 1.081 (0.830-1.407) |

### How did incidence rates and prevalence vary by demographic factors before and after adjustment for age, sex, and BMI (OSA only) in 2019?

Incidence Rate Ratios (IRR) and Prevalence Rate (PR) Ratios for OSA and narcolepsy, by demographic factors, are described in Figures 2 and 3. Incidence and prevalence of OSA was lower among females than males before and after adjustment for age (age-adjusted IRR 0.56, 95% CI 0.54-0.57; age-adjusted PR 0.42, 95% CI 0.41-0.42), with differences increasing after adjustment for BMI (age-BMI-adjusted IRR 0.43, 95% CI 0.42-0.45; age-BMI-adjusted PR 0.34, 95% CI 0.34-0.35). After adjustment for age and sex, OSA incidence rates and prevalence were lower in Other and South Asian ethnic groups compared to White people, lower in rural areas compared to urban areas, and higher in areas of higher deprivation compared to least deprived areas. After further adjustment for BMI, compared to White people, incidence rates and prevalence were higher among South Asian people and lower among Black people; frequency remained lower in rural areas compared to urban areas; rates were lower in the smallest practices compared to other practice size quintiles; and differences by area-based deprivation disappeared.

**Figure 2.**
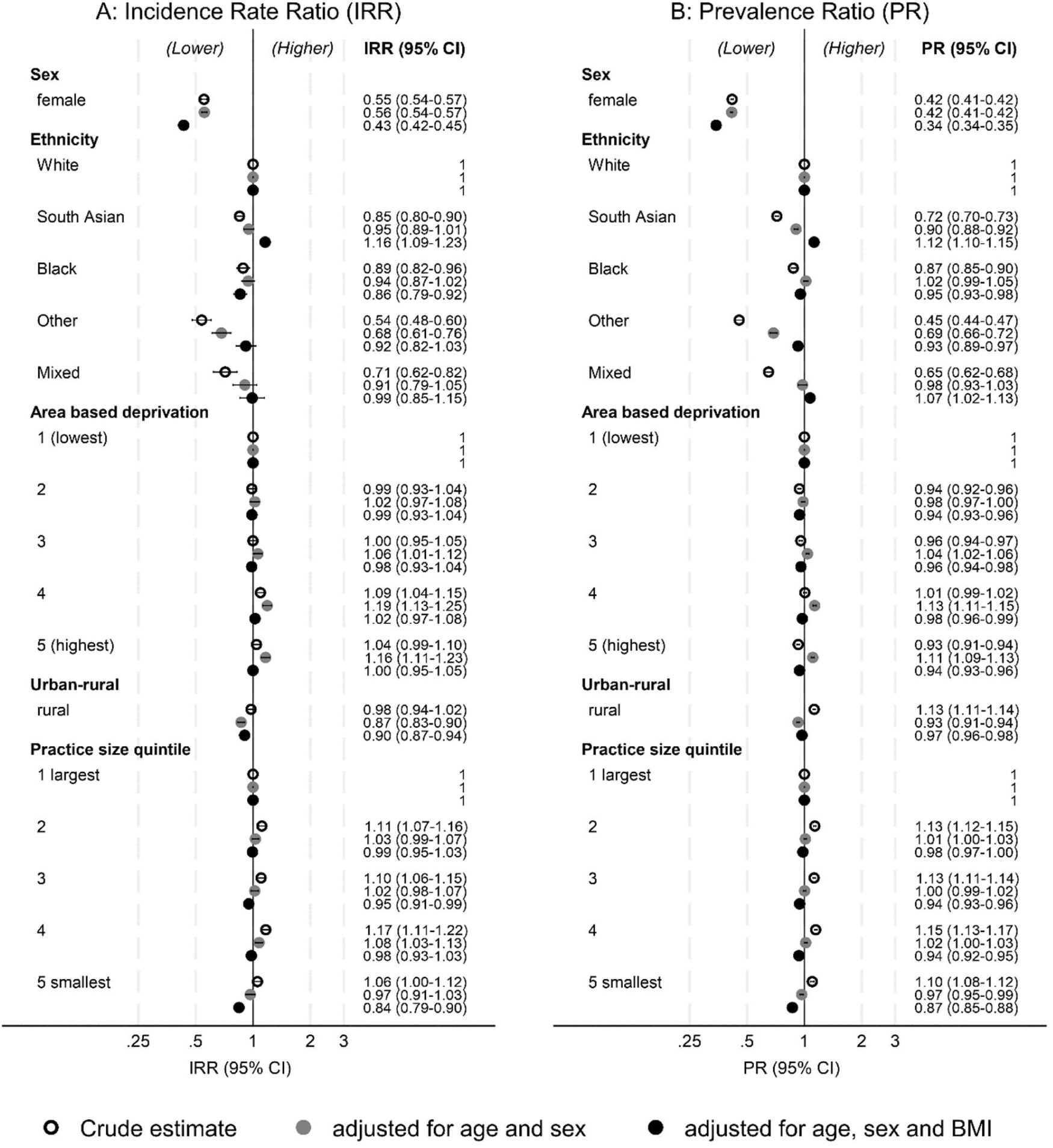
2019 Incidence Rate Ratios and 2019 Prevalence Ratios for diagnosed Obstructive Sleep Apnoea in adults in England

**Figure 3.**
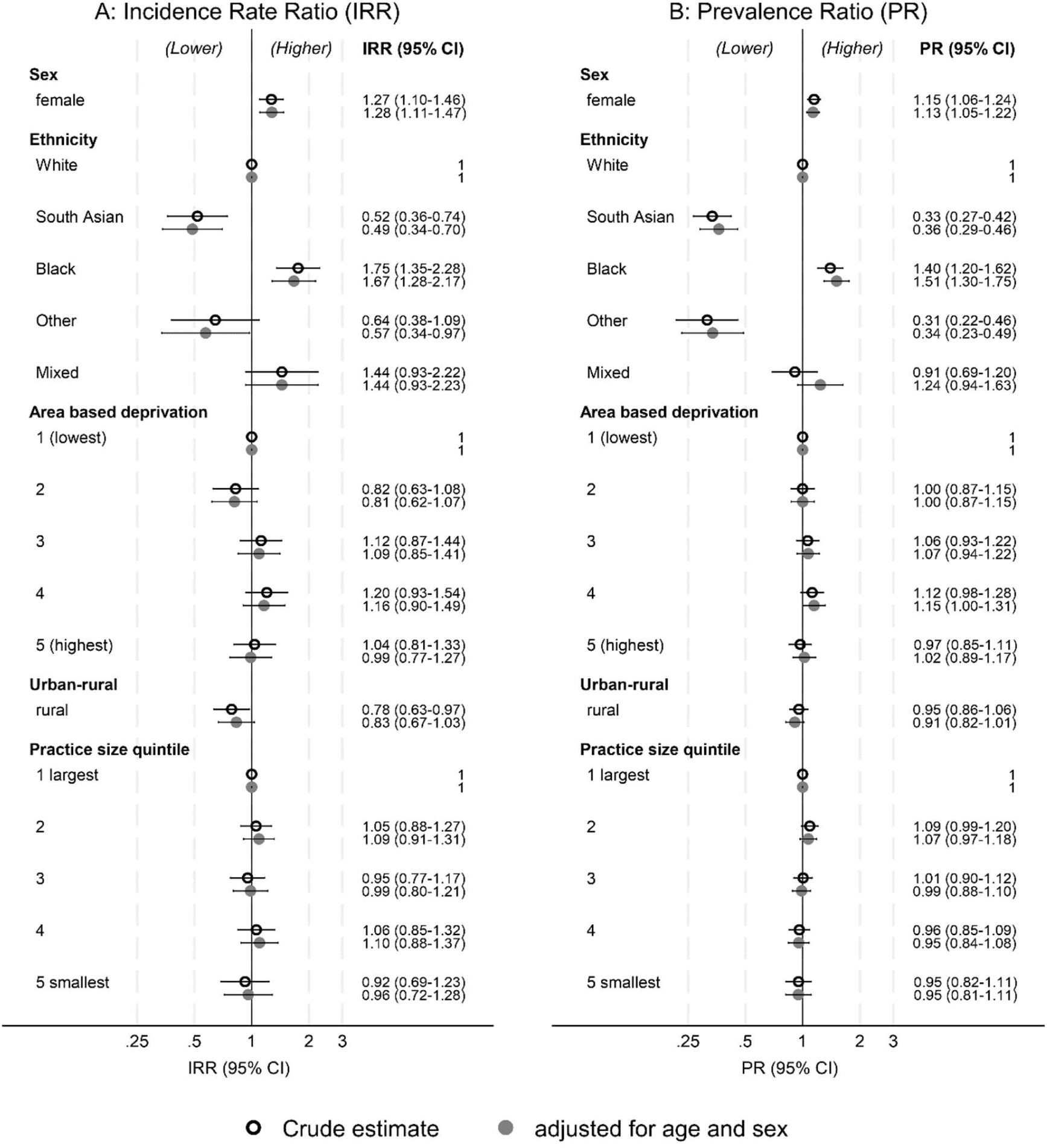
2019 Incidence Rate Ratios and 2015-2019 Prevalence Ratios for diagnosed narcolepsy in England

Crude incidence and prevalence rates of narcolepsy were 27% (95% CI 10%-45%) and 15% (95% CI 5%-24%) higher among women than men. IRRs and PRs described minimal variation by demographic factors before or after adjustment for age and sex. Exceptions were strong statistical evidence of lower incidence and prevalence in South Asians and higher incidence and prevalence in Black people, compared to White people.

### How have incidence rates, prevalence and variation by demographic factors changed over time in England?

Figure 4 and Supplementary Appendix Tables 3 and 4 describe crude diagnosed incidence rates and age-sex standardised diagnosed prevalence between 2000 and 2019. There was an increase in all estimates except for narcolepsy incidence, for which the pattern was less conclusive. There were minimal differences between crude and standardised prevalence rates (Supplementary Appendix table 4). OSA incidence rates multiplied by 6 between 2000 and 2019 (IRR 5.81, 95% CI 5.58-6.06), increases are not associated with changes in population age and sex structures (age, sex adjusted IRR 5.79, 95% CI 5.56-6.04) and only partially associated with increases in OSA incidence rates over time (age, sex, BMI adjusted IRR 3.86, 95% CI 3.68-4.05) Supplementary Figure 3.

**Figure 4:**
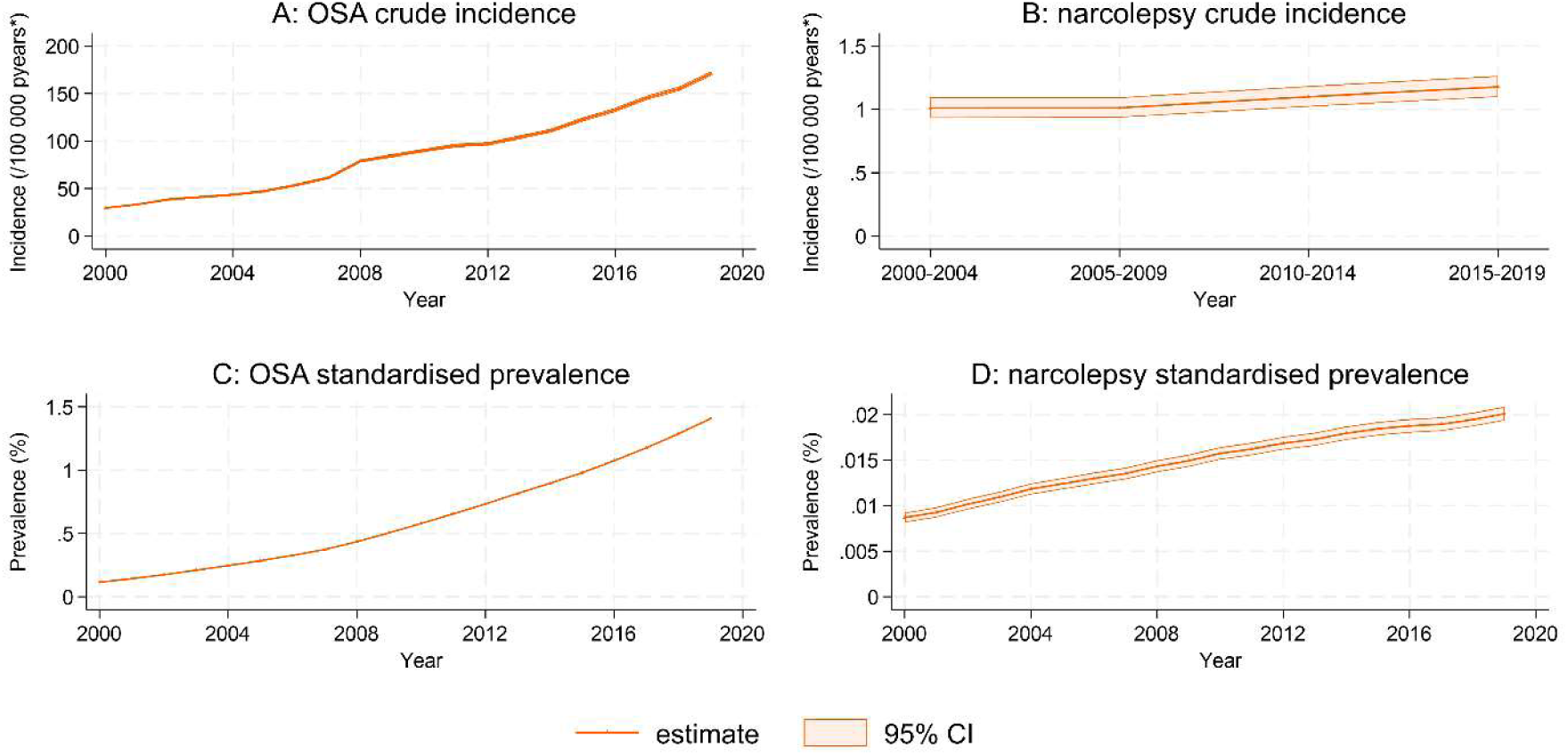
Crude incidence and standardised prevalence of OSA and narcolepsy in England from 2000 to 2019 *pyears = person years

Incidence rates of OSA and narcolepsy by age group and BMI category (OSA only), in 5-year time periods, are described in Figure 5. The peak age group for OSA incidence rates was 45-55 years in 2000-2004, transitioning to 55-65 years by 2015-2019. The age distribution at narcolepsy diagnosis appears to move towards younger people over time.

**Figure 5:**
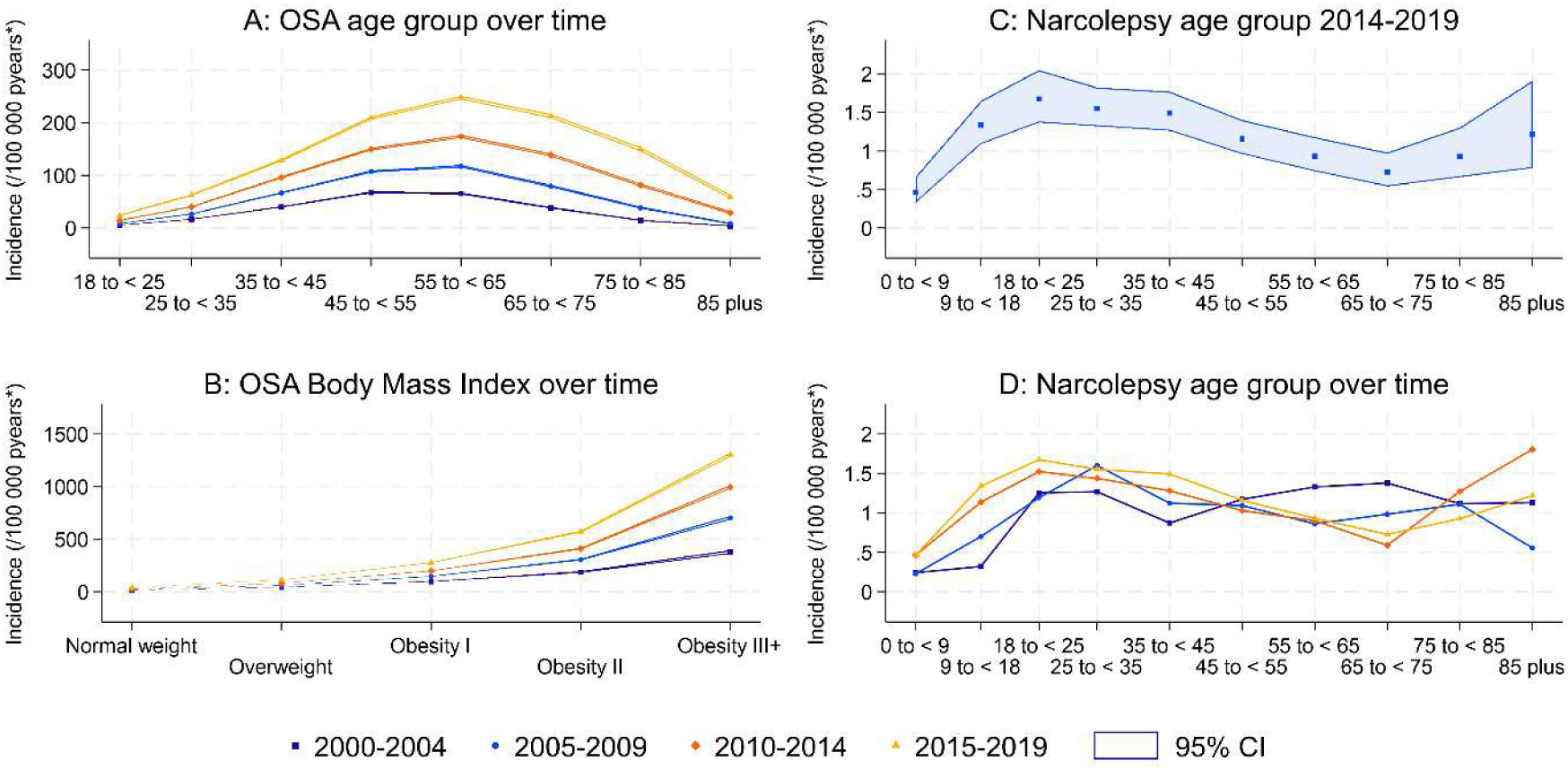
Incidence of Obstructive Sleep Apnoea (OSA) and narcolepsy in England by age group and Body Mass Index (OSA only) stratified by 5-year calendar year categories Graph C does not include 95% confidence intervals as these are wide and overlapping.

Incidence rate ratios for OSA and narcolepsy in each 5-year time period, adjusted for age, sex and BMI (OSA only), are described in Supplementary Appendix Figure 4 and 5, respectively. The association between OSA incidence and sex decreased over time. While people of South Asian ethnicity had consistently elevated incidence rates of OSA compared to White people from 2005-2009 to 2015-2019, the magnitude and direction of IRRs vary for Black and Mixed ethnic groups. Patterns of variation in incidence rates for all other variables were consistent over time, especially from 2005-2009 to 2015-2019.

### Secondary analyses

The primary care study population included 303,926 prevalent and 219,470 incident cases of OSA, and 8,383 prevalent and 3,675 incident cases of narcolepsy. Crude incidence rates and standardised prevalence by time and country in the primary care only analyses are provided in Supplementary Appendix Tables 5-8, Supplementary Appendix Figure 7). OSA prevalence and incidence rates increased over time in all UK nations, increases were greatest in Northern Ireland and lowest in Scotland. In 2019, incidence rates for OSA were 2.5% (95% CI 2.3-2.8) higher in N Ireland and 1.4% (95% CI 1.3-1.5) higher in Wales, compared with England, after adjusting for age, sex and BMI; rates in Scotland were similar to England. 2019 OSA prevalence was approximately 20% lower in the devolved nations compared to England (Figure 6). There was weak evidence that narcolepsy incidence rates decreased in Northern Ireland from 2000 to 2019 contributing to reduced prevalence in recent years (Supplementary Appendix Figure 7). In 2019, narcolepsy incidence rates and prevalence were lower in Northern Ireland than England, differences were not explained by age and sex differences in population structure (Figure 6). Narcolepsy prevalence appeared to increase in Scotland and Wales from 2000 to 2019 while incidence remained stable.

**Figure 6:**
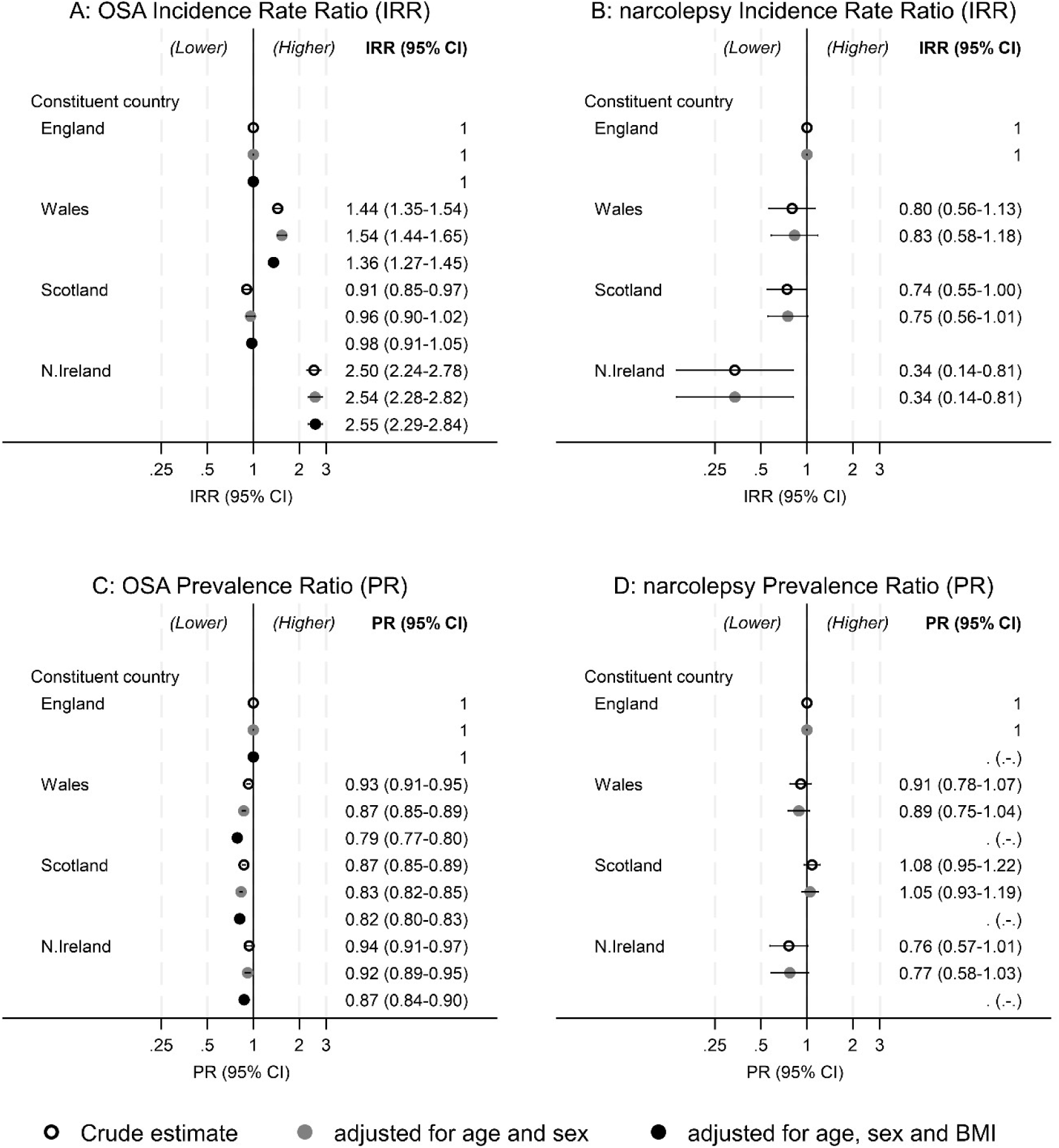
2019 Prevalence Ratios and 2019 (2015-2019 narcolepsy) incidence rate ratios by country in the primary care only analysis population N.Ireland = Northern Ireland. Number of practices: England (1701), Wales (120), Scotland (225), Northern Ireland (54)

### Sensitivity analyses

Crude incidence rates and standardised prevalence estimates using primary care data only were similar in the UK as a whole compared to England, but lower than rates estimated using linked data. Patterns were similar over time for OSA whereas divergence between linked and primary care data only increased over time for narcolepsy (Supplementary Appendix Figure 6). Prevalence ratios and incidence rate ratios were similar in the main linked analysis and primary care only sensitivity analysis for all characteristics (Supplementary Appendix Tables 6-9). Age-sex adjusted IRRs and PRs were similar in the full cohort and cohort restricted to people with BMI measurements (Supplementary Appendix table 10).

## DISCUSSION

### Principal findings

We have provided comprehensive estimates of diagnosed UK prevalence and incidence of OSA and narcolepsy over time, and unique insights concerning variation between demographic subgroups. While diagnosed prevalence increased six-fold for OSA and two-fold for narcolepsy between 2000 and 2019, these remained lower than published estimates of symptomatic UK prevalence(5,6). We confirmed strong associations between diagnosed OSA frequency and key known risk factors (sex, age and BMI) that reduced over time, and uncovered positive associations with South Asian ethnicity, larger practices, urban living, higher area based deprivation, and living in England (prevalence only). With the exception of deprivation, these associations were not explained by differences in age, sex and BMI. As expected, narcolepsy incidence rates peaked in early adulthood. Our findings suggest that narcolepsy diagnosis may be more common in women than men, lowest in South Asians and highest in Black people, and lower in Northern Ireland compared to England.

### Strengths and weaknesses

We have used routinely collected NHS data that represents 25% of the UK population and is representative of the population by age, sex, area-based deprivation and ethnicity (17,21,28,29), reflecting near complete registration of the population in NHS GP practices. However, practices that contribute to CPRD do not accurately represent regions within England due to uneven use of the primary care software systems between regions and clustering within regions(17,28,30); our sample therefore represents small numbers of practices in multiple regions, preventing us from analysing data by region.

Our GP questionnaire validation study indicated that a high proportion (75.3%, 95% CI 69.2-81.3) of OSA cases identified using our primary algorithm were the result of specialist diagnoses and that recorded diagnoses dates were accurate(8). This is a key strength of our study. In contrast, the key weakness of our study is that a moderate proportion 65.2% (95% CI 57.0-73.4) of identified narcolepsy cases are the result of specialist diagnoses and narcolepsy diagnoses were commonly coded years after the specialist diagnosis date. Using our sensitivity analysis algorithm (primary care data only), 85.3% (95% CI 77.3-91.4) of OSA and 71.0% (95% CI 58.8-81.3) of narcolepsy cases were confirmed. These higher proportions of confirmed cases may reflect a lack of accessible information in the GP record regarding hospital diagnoses rather than a true difference. Our observation of similar prevalence ratios and IRRs in our main analysis and sensitivity analysis using the primary care algorithm supports this hypothesis. There are no available estimates of the sensitivity of our algorithms (i.e. the proportion of true diagnosed cases identified) and how this varies by patient characteristics. However, the primary care only algorithm failed to ascertain 14.7% (OSA) and 16.9% (narcolepsy) of gold standard cases from our primary algorithm indicating that not all cases are recorded in primary care data. We may therefore have under- or over-estimated incidence rates and prevalence overall and in specific groups, and the observed increase in diagnoses over time may partially reflect improved recording in primary care and HES data and increasing awareness of sleep disorders. Delayed recording of narcolepsy diagnoses reduces the sensitivity of our narcolepsy prevalence estimates and leads to substantial measurement error in our estimates of narcolepsy incidence rates and rate ratios. For example, the true age distribution at diagnosis is likely to be lower than our estimates.

We have included the devolved nations of the UK, for which linked HES data is not available, in a secondary analysis. These analyses may be affected by practice clustering. We have not attempted to differentiate between narcolepsy types or OSA severity as these data are largely incomplete.

We have described variation in prevalence and incidence of OSA by multiple demographic variables. By combining HES and primary care data, we have largely complete and valid ethnicity data (29) and have measured area-based deprivation and rural urban status. These measures are proxies for individual socio-economic status and rural-urban living and do not account for variation within geographic areas. Ethnicity data were missing for <=2.1% of sleep disorder cases and <=8.1% of the full denominator populations. As data is more complete for people who use GP and hospital services more frequently, our complete case ethnicity PRs and IRR analyses will exclude healthier people; this may bias the findings if the association between ethnicity and sleep disorder frequency is substantially modified by health status. Our time-updated BMI measurements in the OSA cohort use the last BMI measure carried forward; this approach maximises completeness of BMI data but slightly underestimates population level BMI(31). Although missingness was greater in the denominator population (<=16.9%) than people with OSA (<=4.5%), age-sex adjusted ratios were similar in the full population and complete case BMI population. Our complete case approach for characteristics other than BMI will only bias the analysis if the independent effect of BMI in the BMI-adjusted associations differs between people with and without BMI measurements. Our complete case analysis for BMI will be biased if the association between BMI and OSA is different in people with and without missing data.

### Strengths and weaknesses in relation to other studies

To our knowledge, the only previous estimate of prevalence of diagnosed OSA in the UK is from a cross-sectional study using primary care data(12). While that study was focused on people with diabetes, the authors estimated diagnosed prevalence in the full adult population in 2016 to be 0.7% (no CIs reported). This is lower than our 2016 estimate using primary care data only of 0.85% (95% CI 0.83-0.84), potentially reflecting methodological differences (e.g. different code lists for OSA). We estimated the incidence rate of diagnosed narcolepsy per 100,000 person years (95% CI) in England between 2010-2014 and 2015-2019 to be 1.10 (1.02-1.18) and 1.18 (1.10-1.27), respectively. This is comparable to previous estimates using primary care data, which ranged from 1.02 (0.93-1.11) in 2000/2010 (9) to 1.1 (1.0–1.1) in 2003/2014 (11). Our finding of higher incidence in adults than children is also consistent with these studies. Previous studies cited here were affected by the same limitations regarding recording of narcolepsy diagnoses in UK routinely collected data sources as our study, although these were not known at the time.

We have improved understanding of the frequency and distribution of diagnosed OSA and narcolepsy in the UK by, for the first time, describing diagnosed incidence of OSA and diagnosed prevalence of narcolepsy. While age, male sex and BMI have been clearly identified as risk factors for OSA (16), further characterisation of the OSA population is rare. The majority of studies of associations with socio-economic status and deprivation have been conducted in the USA(13).

Findings were heterogeneous with some evidence of increased risk of OSA with lower socioeconomic status that often disappeared after controlling for age, sex and BMI, but little evidence of associations with ethnicity. Mean onset of onset of narcolepsy symptoms was estimated to be 24 years based of sleep disorder centre cohorts from two hospitals in France and Quebec, 10 years lower than our estimates for diagnosed prevalence in the UK(32). We are not aware of any studies comparing narcolepsy incidence and prevalence by wider demographic characteristics(32,33). Our analyses therefore represent the best available data on this topic, despite known limitations.

### Meaning of the study: possible mechanisms and implications for clinicians or policymakers

In England, approximately 1.5% of the adult population have been diagnosed with OSA and 0.02% of the full population have a recorded diagnosis of narcolepsy in routinely collected UK population-based health data. Diagnosis rates of OSA increased approximately 6-fold between 2000 and 2019; and there is some evidence of a smaller increase in narcolepsy diagnosis rates in children and young adults leading to a doubling of diagnosed narcolepsy prevalence. These increases are likely to be due to increasing awareness and capacity to refer for investigations, diagnose and treat these conditions. For example, NICE guidelines for OSA management were developed in 2008(14), the British Lung Foundation published a toolkit for commissioning and planning local services in 2015(34), and awareness of narcolepsy in children increased when cases were associated with the Pandemrix vaccine for H1N1 influenza in 2009(9). The increased incidence of OSA is also partly associated with an aging and increasingly obese population. The increased incidence of narcolepsy in younger age groups may be associated with incorrect diagnoses following increases in awareness but not knowledge. Diagnosed prevalence remains lower than the predicted estimate of 4.8% for moderate to severe OSA based on prevalence estimates from geographically close countries with similar age, sex, BMI and ethnicity distributions (5), and a general population questionnaire study estimate of 0.047% (95% CI 0.016-0.078) for narcolepsy (6). This supports calls to increase medical education and the capacity to identify, investigate, diagnose and treat sleep disorders (16,35).

We found that male sex, obesity and older age are strongly associated with OSA diagnoses. However, in line with a growing consensus that OSA involves a clinical spectrum extending beyond these traditionally recognised risk factors(36), diagnosis is not uncommon, and increasing, in women, younger people and people who are categorised as normal- or over-weight. Multiple studies have shown that social and financial inequalities lead to higher levels of obesity in more deprived areas, with negative implications for life expectancy and health(37). Our observation of elevated rates of diagnosed OSA in areas of high deprivation, that attenuate after adjustment for BMI, suggest that OSA may be a further consequence of poverty-linked inequalities. Our finding of lower rates of OSA and narcolepsy diagnoses in rural areas and OSA in the smallest practices may be associated with access to care(16). Lack of access to care may also contribute to the low rates in Northern Ireland. Persistently strong associations with ethnicity were largely explained by age, sex and BMI for OSA but not by age and sex for narcolepsy.

### Unanswered questions and future research

Variation in the diagnosed frequency of OSA and narcolepsy reflects a combination of the symptomatic frequency of these conditions, inequalities in access to care, and differences in awareness and perceptions of sleep disorders and their symptoms. We have not attempted to understand the influence of these factors, beyond the role of age, sex and obesity in each association.

Future studies of OSA and narcolepsy frequency should consider the challenges revealed by systematic reviews of studies measuring narcolepsy and symptomatic OSA frequency(3–5,33). These systematic reviews reveal widespread heterogeneity in estimates due to methodological variation and limitations of individual study methodologies and data sources. To our knowledge, none of these studies have described demographic variation in the diagnosed frequency of OSA and narcolepsy in detail, preventing international comparisons with our analysis. Future international studies using routinely collected data should be supported by robust assessment of the validity of OSA and narcolepsy measurements; in the UK direct collection and curation of diagnoses in hospital outpatient data are needed to provide better estimates of narcolepsy incidence and prevalence(8,38).

There is a need for further quantitative and qualitative work to more comprehensively describe the availability of sleep medicines services, geographic variation in diagnosed incidence and prevalence, and how societal attitudes to sleep drive differential presentation and diagnosis.

## Conclusion

Diagnosed UK prevalence of OSA and narcolepsy are lower than expected despite increases over time and the substantial impact of these conditions on health, fulfilment and economic wellbeing(34,39). High quality data and research is needed to inform sleep disorder education and health policy initiatives that increase diagnosis rates to expected levels and reduce unexplained variation between demographic groups.

## Data Availability

This study is based in part on data from the Clinical Practice Research Datalink obtained under licence from the UK Medicines and Healthcare products Regulatory Agency. The terms of our licence to access the data preclude us from sharing individual patient data with third parties. The raw data may be requested directly from CPRD following their usual procedures.

## Acknowledgements

This study is based in part on data from the Clinical Practice Research Datalink obtained under licence from the UK Medicines and Healthcare products Regulatory Agency. Hospital Episodes Statistics Data and Office of National Statistics Mortality data were reused with the permission of The Health & Social Care Information Centre (All rights reserved, Copyright (2021)). The data are provided by patients and collected by the NHS as part of their care and support.

## SUPPLEMENTARY APPENDIX

UK incidence and prevalence of Obstructive Sleep Apnoea and narcolepsy: a population-based descriptive study

**SA Figure 1:**
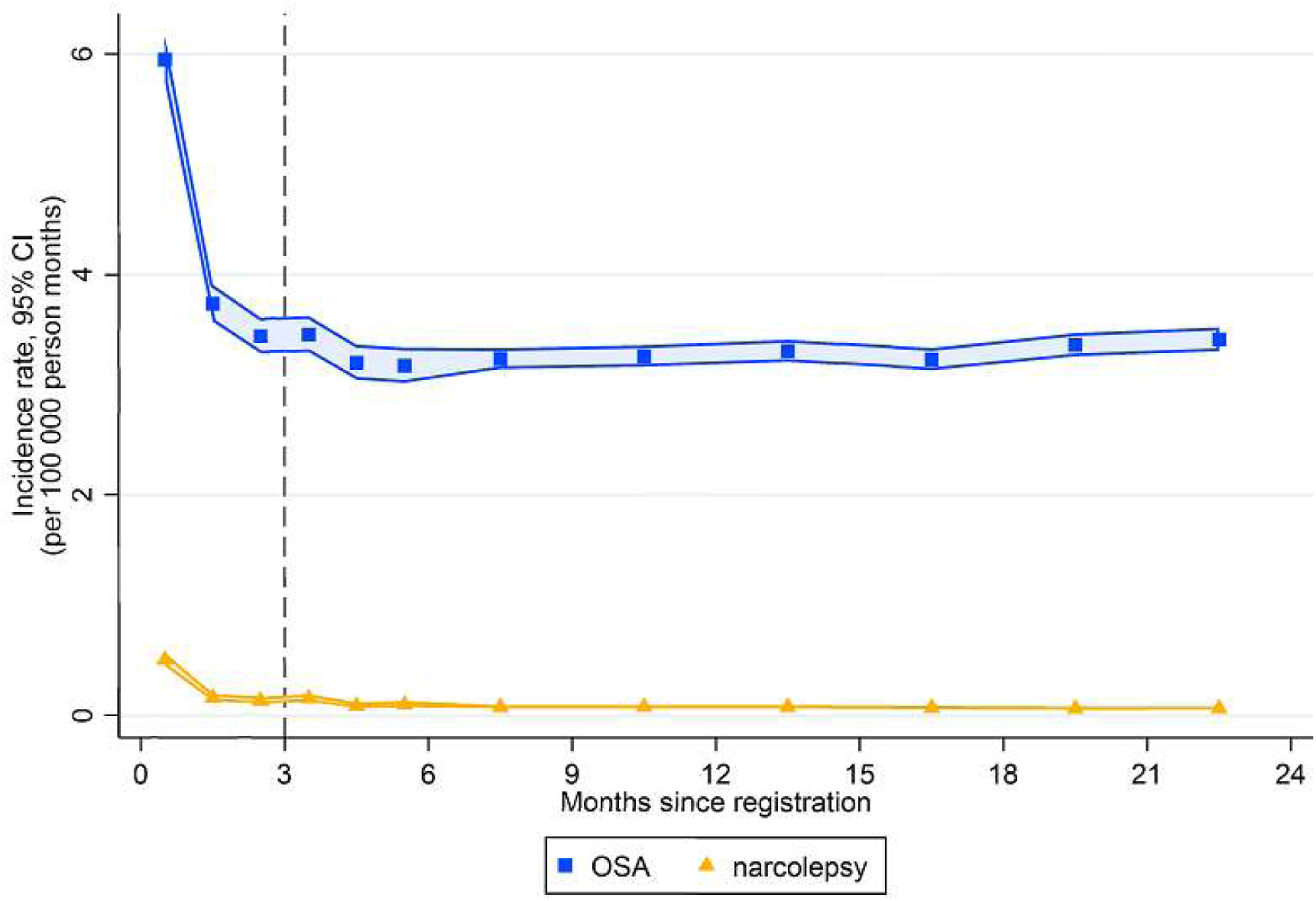
Incidence rate by months since registration in CPRD practice OSA = Obstructive Sleep Apnoea. Black dotted line = 3 months since registration

**SA Figure 2:**
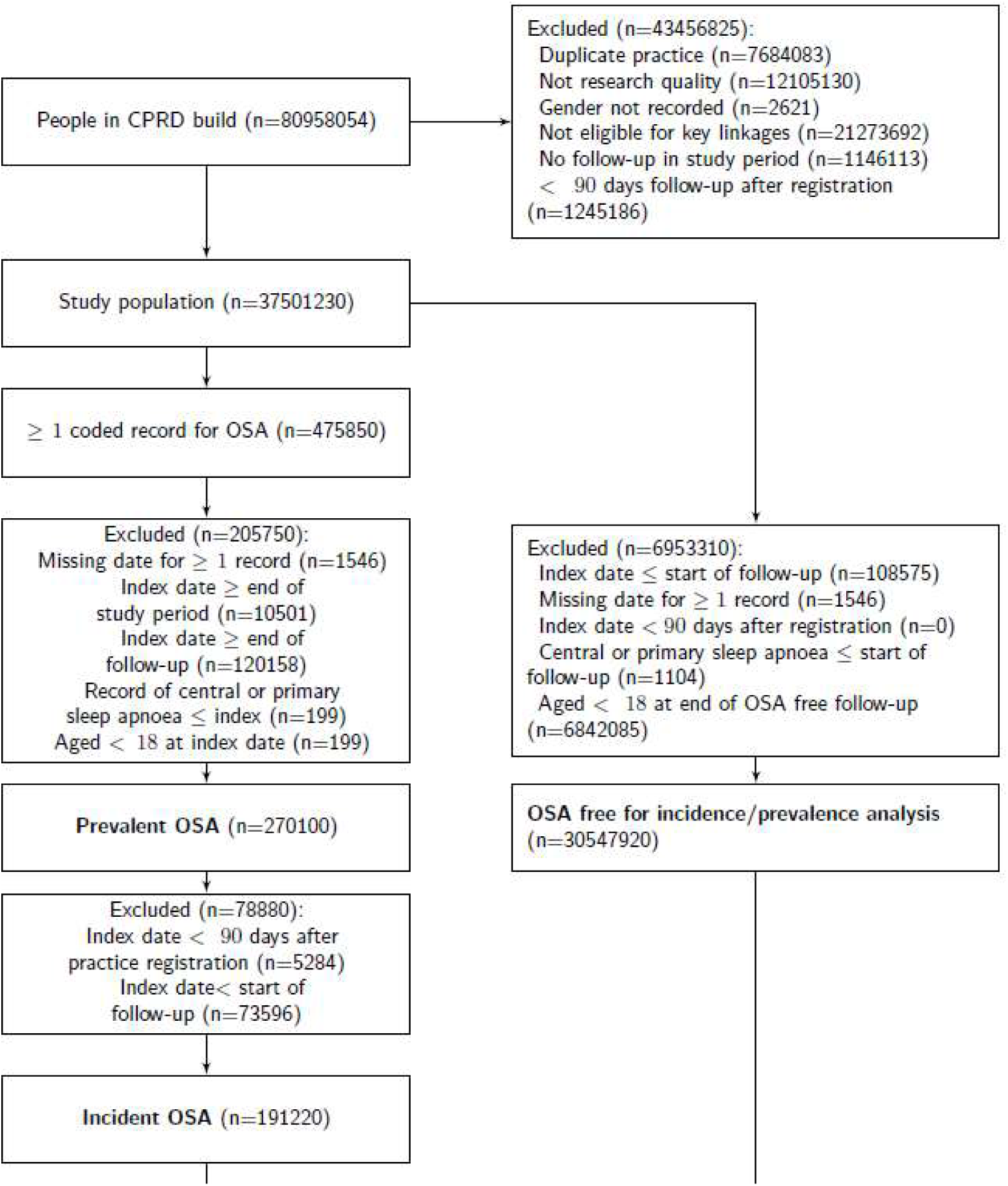
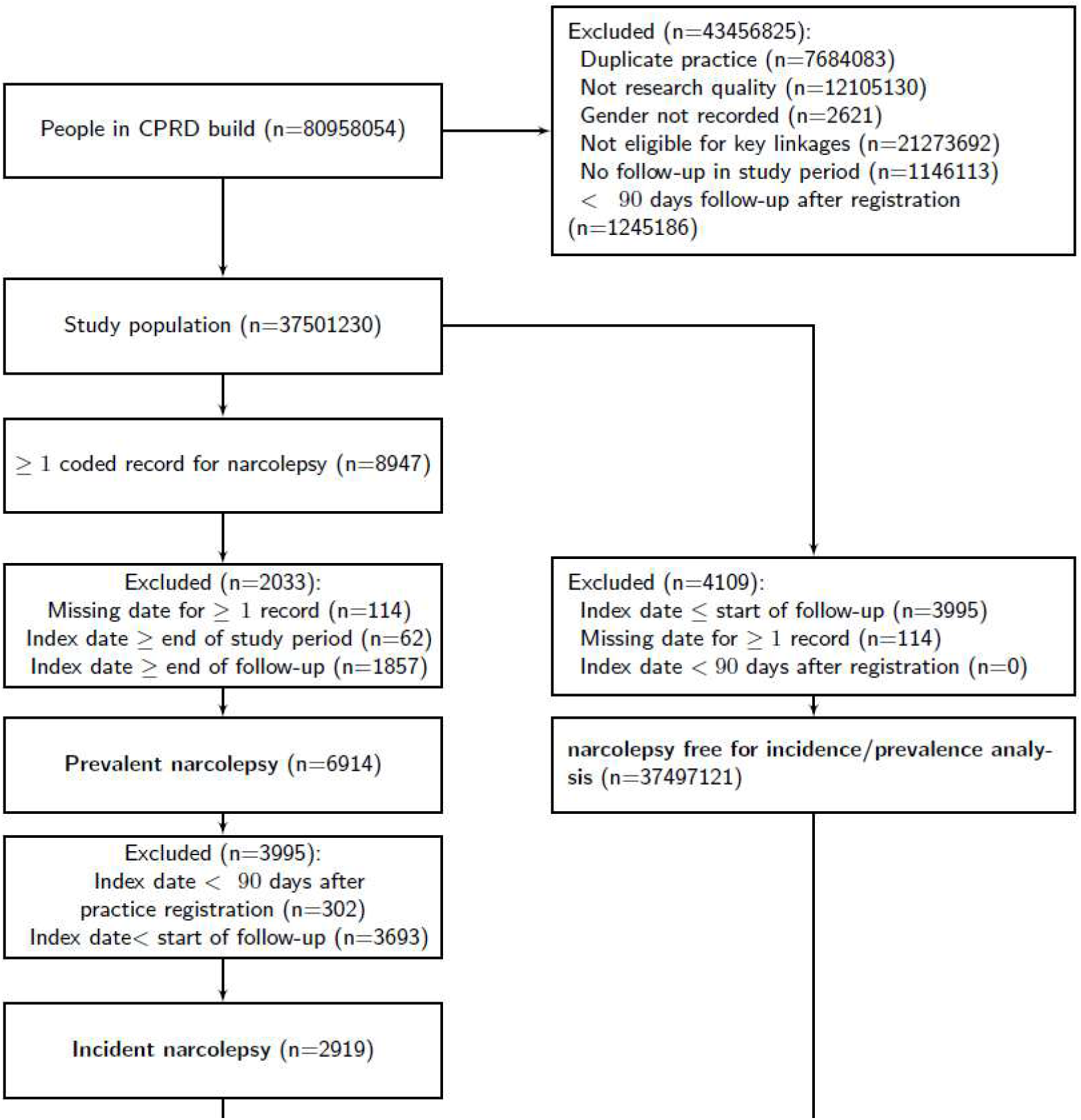
Participant flow charts

**SA Figure 3:**
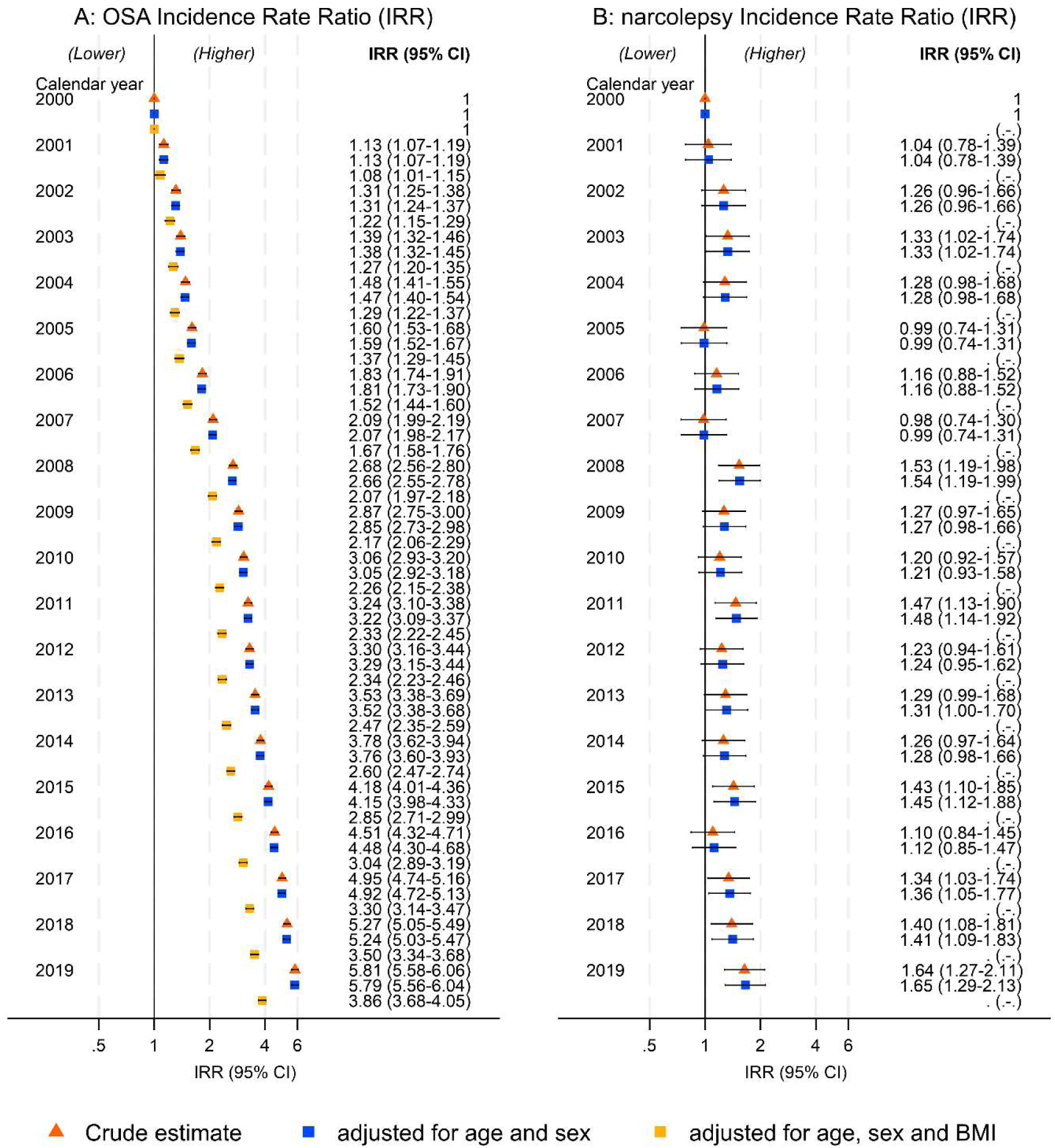
Incidence Rate Ratios by calendar year adjusted for age, sex and BMI (OSA only)

**SA Figure 4:**
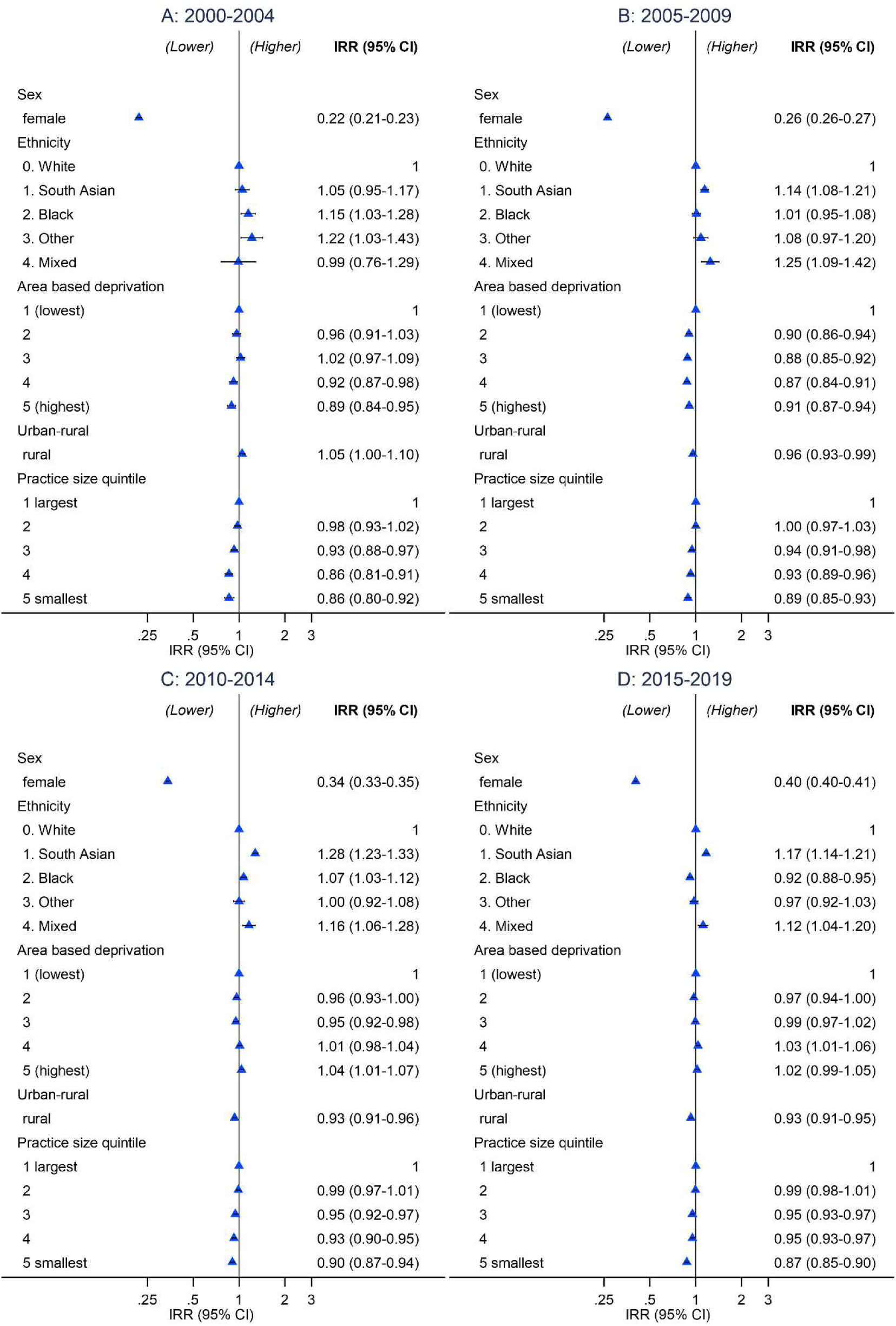
Age, sex and BMI adjusted incident Rate Ratios by demographic characteristics for diagnosed OSA in adults in England from 2000-2019, stratified by 5-year time period.

**SA Figure 5:**
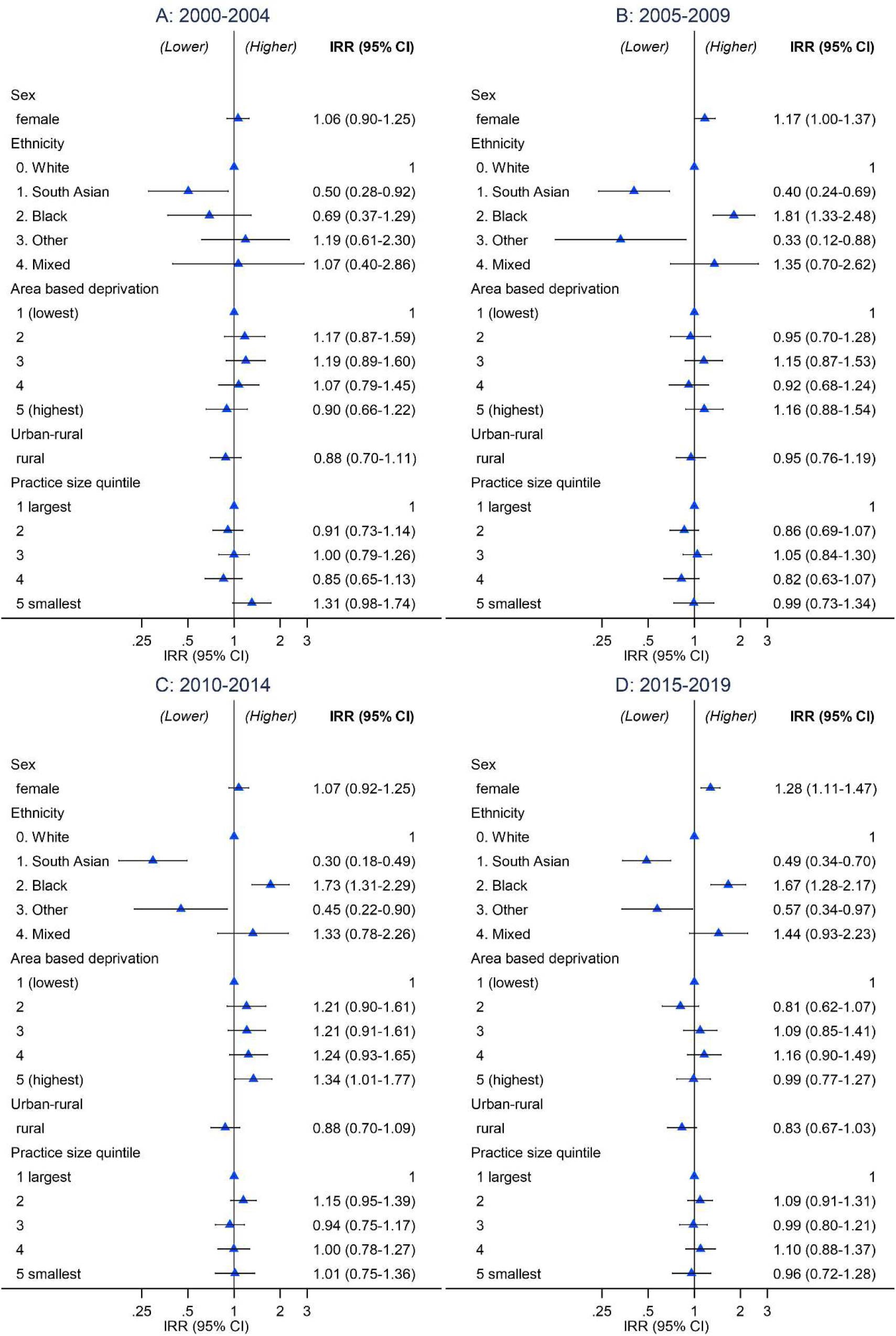
Age and sex adjusted Incident Rate Ratios by demographic characteristics for diagnosed narcolepsy in England from 2000-2019, stratified by 5-year time period.

**SA Figure 6:**
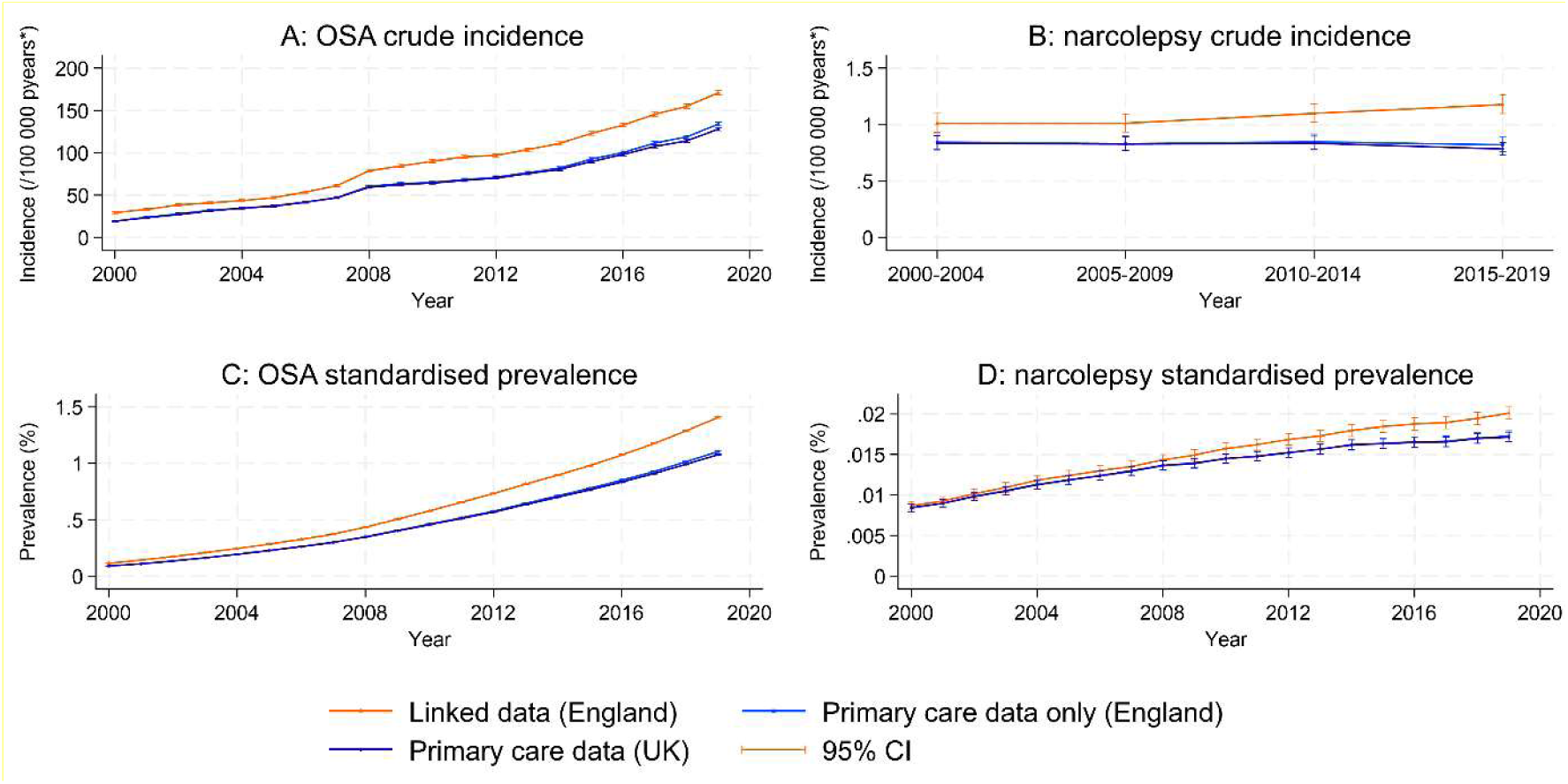
Crude incidence and standardised prevalence of OSA and narcolepsy over time in linked and primary care only data.

**SA Figure 7:**
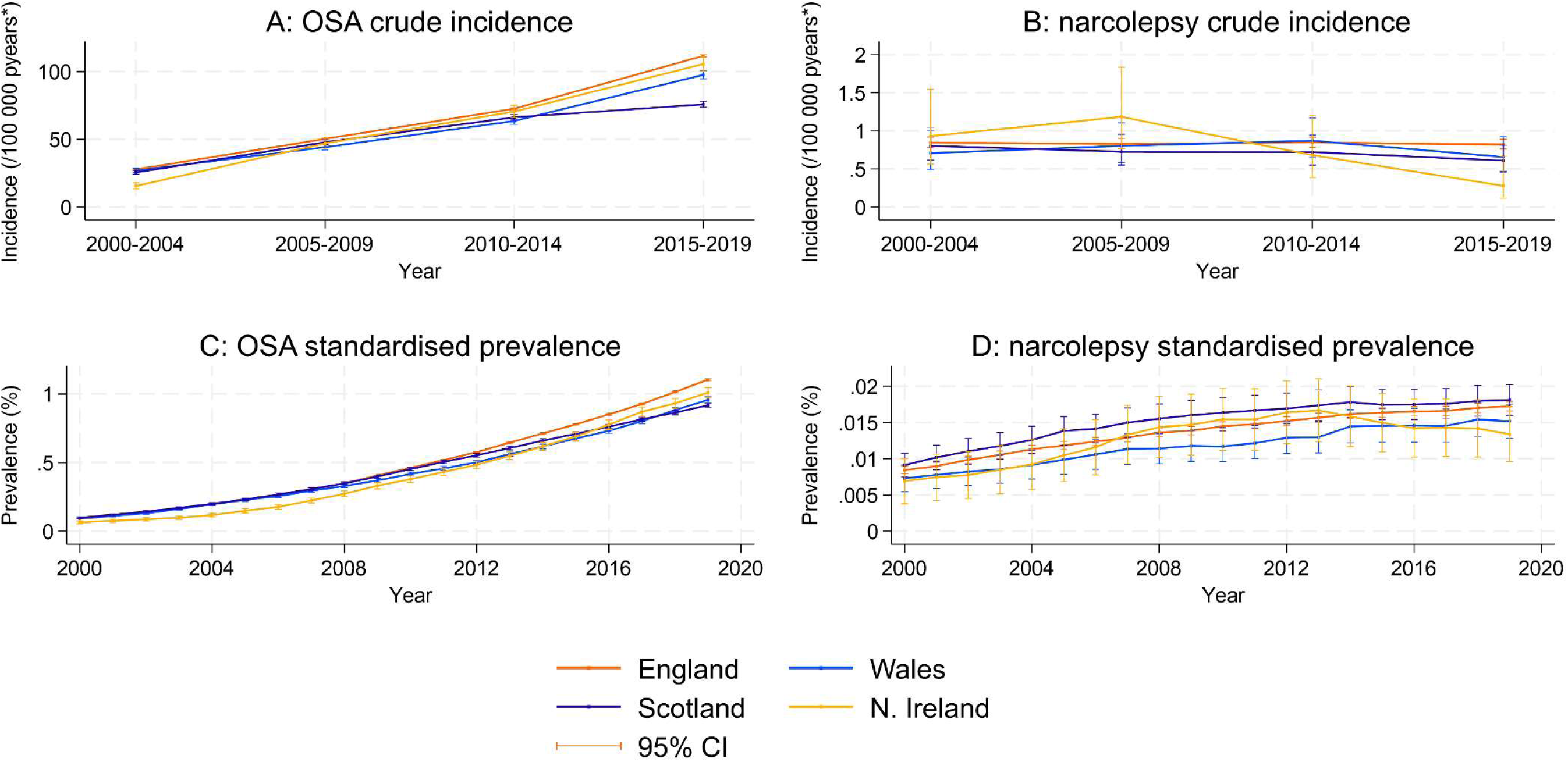
Crude incidence and standardised prevalence of OSA and narcolepsy over time in each UK nation (primary care only data). Caption: Prevalence rates are standardized to the age-sex distribution of each country in each year

---

**SA Table 1:** Data sources, versions and DOIs.

| **Database** | **Version** | **DOI** |
| --- | --- | --- |
| CPRD Aurum | 2022.03.001 | https://doi.org/10.48329/MY9S-4X08 |
| CPRD GOLD | 2022.03.001 | https://doi.org/10.48329/5RHX-ER29 |
| CPRD GOLD HES APC | 2022.01.001 | https://doi.org/10.48329/fagm-ez75 |
| CPRD Aurum HES APC | 2022.01.001 | https://doi.org/10.48329/vagx-9d96 |
| CPRD GOLD ONS deaths | 2022.01.001 | https://doi.org/10.48329/wq7v-x832 |
| CPRD Aurum ONS deaths | 2022.01.001 | https://doi.org/10.48329/Q34F-F505 |
| CPRD GOLD Small Area data (practice) – Carstairs Index | 2022.01.001 | https://doi.org/10.48329/aep7-aq95 |
| CPRD Aurum Small Area data (practice) – Carstairs Index | 2022.01.001 | https://doi.org/10.48329/3saw-rz63 |
| CPRD GOLD Small Area data (patient) – rural-urban classification | 2022.01.001 | https://doi.org/10.48329/y101-0w14 |
| CPRD Aurum Small Area data (patient) – rural-urban classification | 2022.01.001 | https://doi.org/10.48329/aytt-h222 |

**SA Table 2:** Composition of prevalent and incident cases and denominators A: Obstructive Sleep Apnoea (OSA) in adults (2019) B: Narcolepsy in the full population (2019 prevalence, 2014 to 2019 incidence)

| A (OSA) | Prevalent cases | Study population (n) | Incident cases | Person-years (/100 000) |
| --- | --- | --- | --- | --- |
| **Full study population** | 143277 (100.0%) | 10360607 (100.0%) | 17515 (100.0%) | 102.4596 (100.0%) |
| **Age group** |  |  |  |  |
| 18 to < 25 | 694 (0.5%) | 1023006 (9.9%) | 328 (1.9%) | 10.74129 (10.5%) |
| 25 to < 35 | 5737 (4.0%) | 1936833 (18.7%) | 1470 (8.4%) | 19.23007 (18.8%) |
| 35 to < 45 | 15688 (10.9%) | 1821663 (17.6%) | 2756 (15.7%) | 18.00595 (17.6%) |
| 45 to < 55 | 32549 (22.7%) | 1800111 (17.4%) | 4225 (24.1%) | 17.64058 (17.2%) |
| 55 to < 65 | 40265 (28.1%) | 1548317 (14.9%) | 4331 (24.7%) | 15.05144 (14.7%) |
| 65 to < 75 | 31837 (22.2%) | 1182036 (11.4%) | 2880 (16.4%) | 11.48233 (11.2%) |
| 75 to < 85 | 13910 (9.7%) | 734407 (7.1%) | 1291 (7.4%) | 7.194917 (7.0%) |
| 85 plus | 2597 (1.8%) | 314234 (3.0%) | 234 (1.3%) | 3.113063 (3.0%) |
| **Sex** |  |  |  |  |
| male | 101049 (70.5%) | 5176054 (50.0%) | 11232 (64.1%) | 50.92628 (49.7%) |
| female | 42228 (29.5%) | 5184553 (50.0%) | 6283 (35.9%) | 51.53336 (50.3%) |
| **Ethnicity** |  |  |  |  |
| 0. White | 123610 (86.3%) | 7929876 (76.5%) | 14786 (84.4%) | 78.22609 (76.3%) |
| 1. South Asian | 8162 (5.7%) | 730483 (7.1%) | 1166 (6.7%) | 7.262222 (7.1%) |
| 2. Black | 5753 (4.0%) | 421808 (4.1%) | 703 (4.0%) | 4.192815 (4.1%) |
| 3. Other | 2158 (1.5%) | 304707 (2.9%) | 308 (1.8%) | 3.032032 (3.0%) |
| 4. Mixed | 1440 (1.0%) | 142490 (1.4%) | 192 (1.1%) | 1.422215 (1.4%) |
| Missing | 2154 (1.5%) | 831243 (8.0%) | 360 (2.1%) | 8.324272 (8.1%) |
| **Region (number of practices)** |  |  |  |  |
| North East (52) | 5655 (3.9%) | 318806 (3.1%) | 543 (3.1%) | 3.190468 (3.1%) |
| North West (309) | 24223 (16.9%) | 1913401 (18.5%) | 3216 (18.4%) | 18.94195 (18.5%) |
| Yorks & The Humber (41) | 4955 (3.5%) | 360693 (3.5%) | 566 (3.2%) | 3.57356 (3.5%) |
| East Midlands (35) | 3044 (2.1%) | 242177 (2.3%) | 334 (1.9%) | 2.397704 (2.3%) |
| West Midlands (277) | 24010 (16.8%) | 1643109 (15.9%) | 3006 (17.2%) | 16.23146 (15.8%) |
| East of England (44) | 6508 (4.5%) | 413357 (4.0%) | 591 (3.4%) | 4.072668 (4.0%) |
| London (318) | 25672 (17.9%) | 2160670 (20.9%) | 3225 (18.4%) | 21.44683 (20.9%) |
| South East (239) | 30567 (21.3%) | 2076796 (20.0%) | 3817 (21.8%) | 20.60352 (20.1%) |
| South West (145) | 18643 (13.0%) | 1231598 (11.9%) | 2217 (12.7%) | 12.00149 (11.7%) |
| **Area based deprivation** |  |  |  |  |
| 1 (least deprived) | 18673 (13.0%) | 1299876 (12.5%) | 2144 (12.2%) | 12.88049 (12.6%) |
| 2 | 27961 (19.5%) | 2067191 (20.0%) | 3372 (19.3%) | 20.54984 (20.1%) |
| 3 | 32279 (22.5%) | 2348653 (22.7%) | 3851 (22.0%) | 23.1538 (22.6%) |
| 4 | 31323 (21.9%) | 2166843 (20.9%) | 3884 (22.2%) | 21.32207 (20.8%) |
| 5 (most deprived) | 33041 (23.1%) | 2478044 (23.9%) | 4264 (24.3%) | 24.55345 (24.0%) |
| **Urban-rural** |  |  |  |  |
| urban | 119129 (83.1%) | 8778308 (84.7%) | 14908 (85.1%) | 86.88827 (84.8%) |
| rural | 24034 (16.8%) | 1572456 (15.2%) | 2593 (14.8%) | 15.47453 (15.1%) |
| Missing | 114 (0.1%) | 9843 (0.1%) | 14 (0.1%) | .0968552 (0.1%) |
| **Practice size quintile** |  |  |  |  |
| 1 largest | 50437 (35.2%) | 3941366 (38.0%) | 6225 (35.5%) | 38.99069 (38.1%) |
| 2 | 35019 (24.4%) | 2411357 (23.3%) | 4234 (24.2%) | 23.82257 (23.3%) |
| 3 | 26774 (18.7%) | 1859351 (17.9%) | 3242 (18.5%) | 18.41168 (18.0%) |
| 4 | 19693 (13.7%) | 1341764 (13.0%) | 2462 (14.1%) | 13.21102 (12.9%) |
| 5 smallest | 11354 (7.9%) | 806769 (7.8%) | 1352 (7.7%) | 8.023686 (7.8%) |
| **BMI category** |  |  |  |  |
| Underweight | 432 (0.3%) | 235733 (2.3%) | . (%) | . (%) |
| Normal weight | 11986 (8.4%) | 3508945 (33.9%) | 1682 (9.6%) | 34.9553 (34.1%) |
| Overweight | 33904 (23.7%) | 2995206 (28.9%) | 3959 (22.6%) | 29.38917 (28.7%) |
| Obesity class I | 38244 (26.7%) | 1401261 (13.5%) | 4275 (24.4%) | 13.45145 (13.1%) |
| Obesity class II | 26695 (18.6%) | 520964 (5.0%) | 3172 (18.1%) | 4.843572 (4.7%) |
| Obesity class III+ | 29091 (20.3%) | 289112 (2.8%) | 3645 (20.8%) | 2.51635 (2.5%) |
| Missing | 2925 (2.0%) | 1409386 (13.6%) | 782 (4.5%) | 17.30381 (16.9%) |

| **B (narcolepsy)** | **Prevalent cases** | **Study population (n)** | **Incident cases** | **Person-years (/100 000)** |
| --- | --- | --- | --- | --- |
| **Full study population** | 2662 (100.0%) | 13159742 (100.0%) | 761 (100.0%) | 645.9674 (100.0%) |
| **Age group** |  |  |  |  |
| 0 to < 9 | 21 (0.8%) | 1270179 (9.7%) | 30 (3.9%) | 64.74413 (10.0%) |
| 9 to < 18 | 108 (4.1%) | 1386937 (10.5%) | 88 (11.6%) | 65.72792 (10.2%) |
| 18 to < 25 | 168 (6.3%) | 1161086 (8.8%) | 94 (12.4%) | 56.1519 (8.7%) |
| 25 to < 35 | 399 (15.0%) | 1939578 (14.7%) | 146 (19.2%) | 94.121 (14.6%) |
| 35 to < 45 | 509 (19.1%) | 1822046 (13.8%) | 133 (17.5%) | 89.08833 (13.8%) |
| 45 to < 55 | 488 (18.3%) | 1800325 (13.7%) | 106 (13.9%) | 91.5315 (14.2%) |
| 55 to < 65 | 364 (13.7%) | 1548575 (11.8%) | 69 (9.1%) | 74.1439 (11.5%) |
| 65 to < 75 | 277 (10.4%) | 1182240 (9.0%) | 43 (5.7%) | 59.23645 (9.2%) |
| 75 to < 85 | 220 (8.3%) | 734519 (5.6%) | 33 (4.3%) | 35.62558 (5.5%) |
| 85 plus | 108 (4.1%) | 314257 (2.4%) | 19 (2.5%) | 15.59666 (2.4%) |
| **Sex** |  |  |  |  |
| male | 1245 (46.8%) | 6611703 (50.2%) | 337 (44.3%) | 324.2886 (50.2%) |
| female | 1417 (53.2%) | 6548039 (49.8%) | 424 (55.7%) | 321.6787 (49.8%) |
| **Ethnicity** |  |  |  |  |
| 0. White | 2261 (84.9%) | 9939848 (75.5%) | 621 (81.6%) | 495.1721 (76.7%) |
| 1. South Asian | 77 (2.9%) | 1011101 (7.7%) | 31 (4.1%) | 47.69589 (7.4%) |
| 2. Black | 186 (7.0%) | 585159 (4.4%) | 62 (8.1%) | 28.18765 (4.4%) |
| 3. Other | 28 (1.1%) | 391117 (3.0%) | 14 (1.8%) | 17.34145 (2.7%) |
| 4. Mixed | 53 (2.0%) | 255972 (1.9%) | 21 (2.8%) | 11.62666 (1.8%) |
| Missing | 57 (2.1%) | 976545 (7.4%) | 12 (1.6%) | 45.94366 (7.1%) |
| **Region** |  |  |  |  |
| North East (52) | 91 (3.4%) | 394775 (3.0%) | 17 (2.2%) | 20.4515 (3.2%) |
| North West (309) | 455 (17.1%) | 2421981 (18.4%) | 136 (17.9%) | 117.2262 (18.1%) |
| Yorks & The Humber (41) | 72 (2.7%) | 445481 (3.4%) | 14 (1.8%) | 22.89745 (3.5%) |
| East Midlands (35) | 58 (2.2%) | 295175 (2.2%) | 19 (2.5%) | 15.44438 (2.4%) |
| West Midlands (277) | 431 (16.2%) | 2095827 (15.9%) | 153 (20.1%) | 103.0279 (15.9%) |
| East of England (44) | 105 (3.9%) | 533414 (4.1%) | 20 (2.6%) | 28.51435 (4.4%) |
| London (318) | 581 (21.8%) | 2773362 (21.1%) | 168 (22.1%) | 130.9392 (20.3%) |
| South East (239) | 560 (21.0%) | 2653194 (20.2%) | 157 (20.6%) | 130.6536 (20.2%) |
| South West (145) | 309 (11.6%) | 1546533 (11.8%) | 77 (10.1%) | 76.8128 (11.9%) |
| **Area based deprivation** |  |  |  |  |
| 1 (least deprived) | 325 (12.2%) | 1658427 (12.6%) | 91 (12.0%) | 80.49815 (12.5%) |
| 2 | 507 (19.0%) | 2588319 (19.7%) | 119 (15.6%) | 127.9654 (19.8%) |
| 3 | 613 (23.0%) | 2945071 (22.4%) | 184 (24.2%) | 145.3312 (22.5%) |
| 4 | 604 (22.7%) | 2747216 (20.9%) | 183 (24.0%) | 134.9633 (20.9%) |
| 5 (most deprived) | 613 (23.0%) | 3220709 (24.5%) | 184 (24.2%) | 157.2094 (24.3%) |
| **Urban-rural** |  |  |  |  |
| urban | 2279 (85.6%) | 1.12e+07 (85.0%) | 667 (87.6%) | 547.0087 (84.7%) |
| rural | 381 (14.3%) | 1959232 (14.9%) | 94 (12.4%) | 98.34284 (15.2%) |
| Missing | 2 (0.1%) | 12076 (0.1%) | 0 (0.0%) | .6158447 (0.1%) |
| **Practice size quintile** |  |  |  |  |
| 1 largest | 989 (37.2%) | 4953260 (37.6%) | 280 (36.8%) | 238.9982 (37.0%) |
| 2 | 669 (25.1%) | 3072493 (23.3%) | 188 (24.7%) | 152.2185 (23.6%) |
| 3 | 474 (17.8%) | 2360250 (17.9%) | 132 (17.3%) | 118.3416 (18.3%) |
| 4 | 332 (12.5%) | 1731268 (13.2%) | 106 (13.9%) | 85.51196 (13.2%) |
| 5 smallest | 198 (7.4%) | 1042471 (7.9%) | 55 (7.2%) | 50.89715 (7.9%) |

**SA Table 3:**
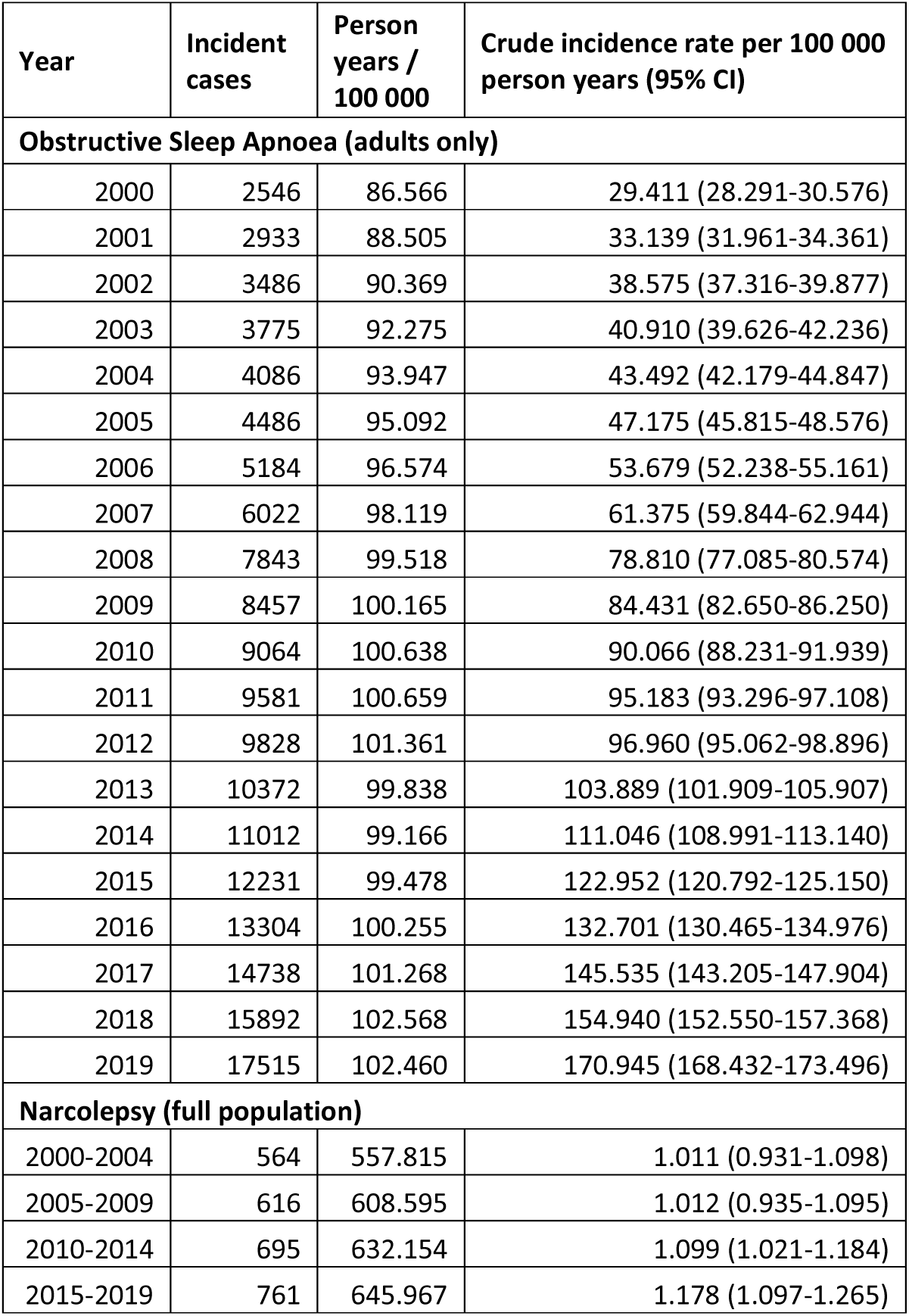
Diagnosed incidence rates for Obstructive Sleep Apneoa (OSA) and narcolepsy in England from 2000 to 2019.

**SA Table 4:** Diagnosed prevalence of Obstructive Sleep Apneoa (OSA) and narcolepsy in England from 2000 to 2019 *directly age/sex standardised to Office of National Statistics population figures for England for each year.

| Year | Prev-<br>alent<br>cases<br>(n) | Study pop-<br>ulation (n) | Crude prevalence<br>% (95% CI) | Standardised*<br>prevalence % (95%<br>CI) | Estimated cases in the<br>population n (95%<br>plausible interval) |
| --- | --- | --- | --- | --- | --- |
| **Obstructive Sleep Apnoea (adults only)** |  |  |  |  |  |
| 2000 | 10305 | 8593444 | 0.120 (0.118-0.122) | 0.116 (0.113-0.118) | 43,959 (43,110-44,807) |
| 2001 | 13012 | 8820998 | 0.148 (0.145-0.150) | 0.142 (0.140-0.145) | 54,522 (53,586-55,458) |
| 2002 | 16223 | 8997943 | 0.180 (0.178-0.183) | 0.174 (0.171-0.177) | 67,125 (66,093-68,157) |
| 2003 | 19905 | 9198929 | 0.216 (0.213-0.219) | 0.209 (0.206-0.212) | 81,260 (80,133-82,388) |
| 2004 | 23738 | 9345873 | 0.254 (0.251-0.257) | 0.246 (0.243-0.249) | 96,091 (94,871-97,312) |
| 2005 | 27820 | 9471842 | 0.294 (0.290-0.297) | 0.285 (0.281-0.288) | 112,392 (111,074-113,711) |
| 2006 | 32444 | 9635545 | 0.337 (0.333-0.340) | 0.327 (0.323-0.331) | 130,319 (128,904-131,734) |
| 2007 | 37566 | 9796975 | 0.383 (0.380-0.387) | 0.373 (0.369-0.377) | 150,075 (148,561-151,588) |
| 2008 | 44147 | 9913300 | 0.445 (0.441-0.449) | 0.434 (0.430-0.438) | 176,396 (174,756-178,036) |
| 2009 | 51623 | 10022265 | 0.515 (0.511-0.520) | 0.504 (0.500-0.509) | 206,620 (204,844-208,396) |
| 2010 | 59348 | 10068216 | 0.589 (0.585-0.594) | 0.579 (0.574-0.583) | 239,370 (237,453-241,288) |
| 2011 | 67161 | 10088116 | 0.666 (0.661-0.671) | 0.655 (0.650-0.660) | 273,710 (271,650-275,769) |
| 2012 | 75317 | 10158121 | 0.741 (0.736-0.747) | 0.733 (0.728-0.738) | 308,408 (306,218-310,598) |
| 2013 | 82248 | 9985639 | 0.824 (0.818-0.829) | 0.816 (0.810-0.821) | 345,579 (343,232-347,925) |
| 2014 | 89893 | 9944074 | 0.904 (0.898-0.910) | 0.896 (0.890-0.902) | 382,838 (380,353-385,323) |
| 2015 | 98502 | 9980677 | 0.987 (0.981-0.993) | 0.979 (0.973-0.985) | 422,103 (419,488-424,719) |
| 2016 | 108061 | 10036444 | 1.077 (1.070-1.083) | 1.074 (1.068-1.081) | 467,120 (464,357-469,882) |
| 2017 | 119347 | 10200979 | 1.170 (1.163-1.177) | 1.175 (1.169-1.182) | 514,299 (511,406-517,191) |
| 2018 | 131873 | 10347122 | 1.274 (1.268-1.281) | 1.287 (1.281-1.294) | 566,746 (563,716-569,777) |
| 2019 | 143277 | 10360607 | 1.383 (1.376-1.390) | 1.406 (1.399-1.414) | 622,528 (619,336-625,720) |
| **Narcolepsy (full population)** |  |  |  |  |  |
| 2000 | 979 | 10649029 | 0.009 (0.009-0.010) | 0.009 (0.008-0.009) | 4,278 (4,009-4,546) |
| 2001 | 1067 | 10935908 | 0.010 (0.009-0.010) | 0.009 (0.009-0.010) | 4,578 (4,303-4,853) |
| 2002 | 1193 | 11157068 | 0.011 (0.010-0.011) | 0.010 (0.010-0.011) | 5,050 (4,763-5,337) |
| 2003 | 1305 | 11413180 | 0.011 (0.011-0.012) | 0.011 (0.010-0.012) | 5,458 (5,161-5,755) |
| 2004 | 1431 | 11605453 | 0.012 (0.012-0.013) | 0.012 (0.011-0.012) | 5,938 (5,629-6,246) |
| 2005 | 1516 | 11777450 | 0.013 (0.012-0.014) | 0.012 (0.012-0.013) | 6,279 (5,962-6,595) |
| 2006 | 1612 | 11992543 | 0.013 (0.013-0.014) | 0.013 (0.012-0.014) | 6,620 (6,297-6,944) |
| 2007 | 1703 | 12208739 | 0.014 (0.013-0.015) | 0.014 (0.013-0.014) | 6,951 (6,620-7,282) |
| 2008 | 1823 | 12375754 | 0.015 (0.014-0.015) | 0.014 (0.014-0.015) | 7,423 (7,081-7,764) |
| 2009 | 1919 | 12533001 | 0.015 (0.015-0.016) | 0.015 (0.014-0.016) | 7,791 (7,442-8,140) |
| 2010 | 2027 | 12606257 | 0.016 (0.015-0.017) | 0.016 (0.015-0.016) | 8,283 (7,922-8,644) |
| 2011 | 2095 | 12668578 | 0.017 (0.016-0.017) | 0.016 (0.016-0.017) | 8,612 (8,242-8,981) |
| 2012 | 2192 | 12784860 | 0.017 (0.016-0.018) | 0.017 (0.016-0.018) | 9,017 (8,639-9,395) |
| 2013 | 2211 | 12605317 | 0.018 (0.017-0.018) | 0.017 (0.017-0.018) | 9,312 (8,924-9,701) |
| 2014 | 2287 | 12585025 | 0.018 (0.017-0.019) | 0.018 (0.017-0.019) | 9,753 (9,353-10,153) |
| 2015 | 2356 | 12646881 | 0.019 (0.018-0.019) | 0.018 (0.018-0.019) | 10,102 (9,693-10,510) |
| 2016 | 2406 | 12736996 | 0.019 (0.018-0.020) | 0.019 (0.018-0.020) | 10,364 (9,949-10,779) |
| 2017 | 2470 | 12959549 | 0.019 (0.018-0.020) | 0.019 (0.018-0.020) | 10,531 (10,115-10,948) |
| 2018 | 2573 | 13146928 | 0.020 (0.019-0.020) | 0.019 (0.019-0.020) | 10,885 (10,463-11,306) |
| 2019 | 2662 | 13159742 | 0.020 (0.019-0.021) | 0.020 (0.019-0.021) | 11,307 (10,876-11,738) |

**SA Table 5:**
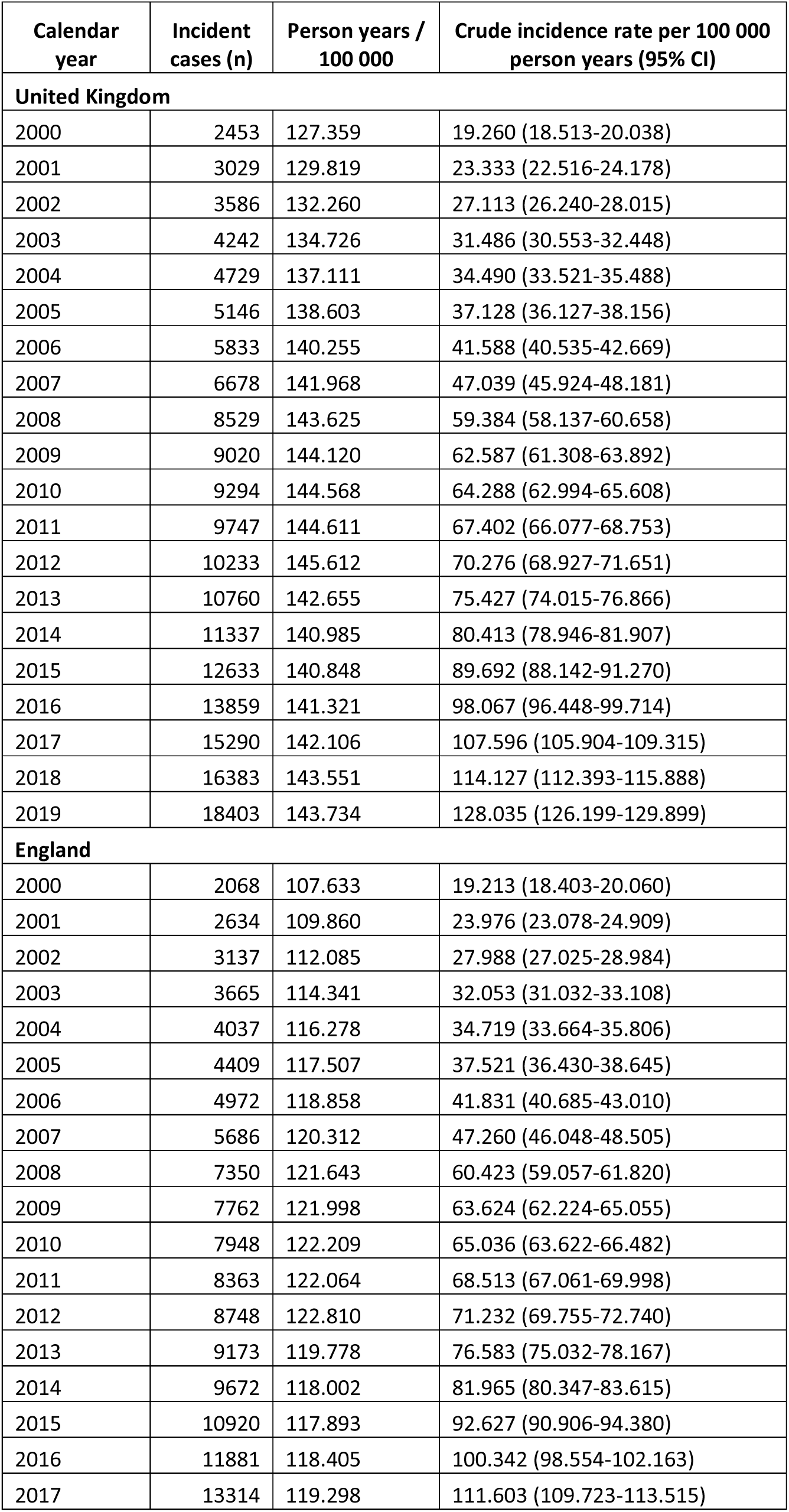
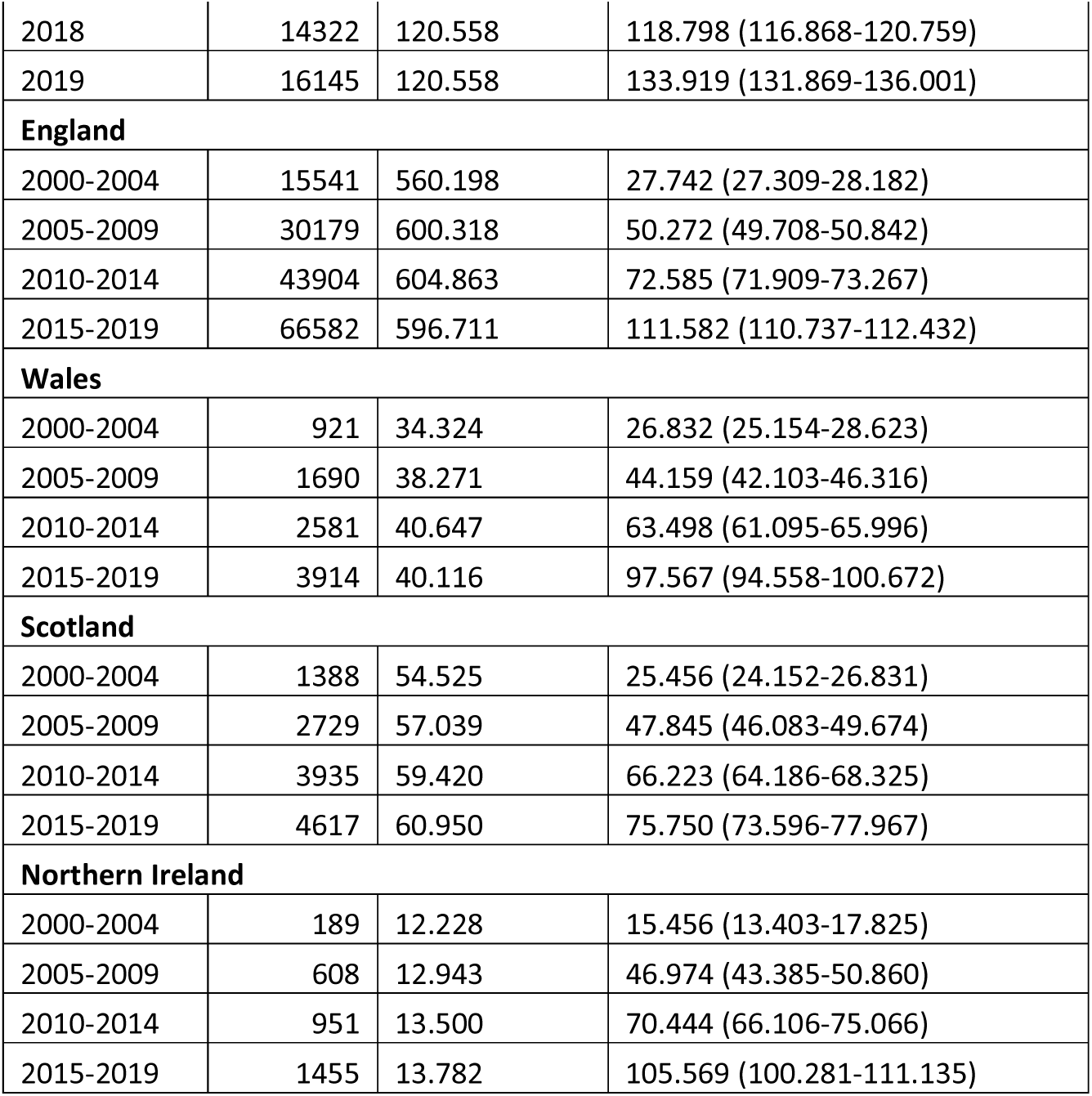
Crude incidence rates by year and country in primary care only study populations (obstructive sleep apnoea - OSA)

| Calendar year | Incident cases (n) | Person years / 100 000 | Crude incidence rate per 100 000 person years (95% CI) |
| --- | --- | --- | --- |
| **United Kingdom** |  |  |  |
| 2000 | 2453 | 127.359 | 19.260 (18.513-20.038) |
| 2001 | 3029 | 129.819 | 23.333 (22.516-24.178) |
| 2002 | 3586 | 132.260 | 27.113 (26.240-28.015) |
| 2003 | 4242 | 134.726 | 31.486 (30.553-32.448) |
| 2004 | 4729 | 137.111 | 34.490 (33.521-35.488) |
| 2005 | 5146 | 138.603 | 37.128 (36.127-38.156) |
| 2006 | 5833 | 140.255 | 41.588 (40.535-42.669) |
| 2007 | 6678 | 141.968 | 47.039 (45.924-48.181) |
| 2008 | 8529 | 143.625 | 59.384 (58.137-60.658) |
| 2009 | 9020 | 144.120 | 62.587 (61.308-63.892) |
| 2010 | 9294 | 144.568 | 64.288 (62.994-65.608) |
| 2011 | 9747 | 144.611 | 67.402 (66.077-68.753) |
| 2012 | 10233 | 145.612 | 70.276 (68.927-71.651) |
| 2013 | 10760 | 142.655 | 75.427 (74.015-76.866) |
| 2014 | 11337 | 140.985 | 80.413 (78.946-81.907) |
| 2015 | 12633 | 140.848 | 89.692 (88.142-91.270) |
| 2016 | 13859 | 141.321 | 98.067 (96.448-99.714) |
| 2017 | 15290 | 142.106 | 107.596 (105.904-109.315) |
| 2018 | 16383 | 143.551 | 114.127 (112.393-115.888) |
| 2019 | 18403 | 143.734 | 128.035 (126.199-129.899) |
| **England** |  |  |  |
| 2000 | 2068 | 107.633 | 19.213 (18.403-20.060) |
| 2001 | 2634 | 109.860 | 23.976 (23.078-24.909) |
| 2002 | 3137 | 112.085 | 27.988 (27.025-28.984) |
| 2003 | 3665 | 114.341 | 32.053 (31.032-33.108) |
| 2004 | 4037 | 116.278 | 34.719 (33.664-35.806) |
| 2005 | 4409 | 117.507 | 37.521 (36.430-38.645) |
| 2006 | 4972 | 118.858 | 41.831 (40.685-43.010) |
| 2007 | 5686 | 120.312 | 47.260 (46.048-48.505) |
| 2008 | 7350 | 121.643 | 60.423 (59.057-61.820) |
| 2009 | 7762 | 121.998 | 63.624 (62.224-65.055) |
| 2010 | 7948 | 122.209 | 65.036 (63.622-66.482) |
| 2011 | 8363 | 122.064 | 68.513 (67.061-69.998) |
| 2012 | 8748 | 122.810 | 71.232 (69.755-72.740) |
| 2013 | 9173 | 119.778 | 76.583 (75.032-78.167) |
| 2014 | 9672 | 118.002 | 81.965 (80.347-83.615) |
| 2015 | 10920 | 117.893 | 92.627 (90.906-94.380) |
| 2016 | 11881 | 118.405 | 100.342 (98.554-102.163) |
| 2017 | 13314 | 119.298 | 111.603 (109.723-113.515) |
| 2018 | 14322 | 120.558 | 118.798 (116.868-120.759) |
| 2019 | 16145 | 120.558 | 133.919 (131.869-136.001) |
| **England** |  |  |  |
| 2000-2004 | 15541 | 560.198 | 27.742 (27.309-28.182) |
| 2005-2009 | 30179 | 600.318 | 50.272 (49.708-50.842) |
| 2010-2014 | 43904 | 604.863 | 72.585 (71.909-73.267) |
| 2015-2019 | 66582 | 596.711 | 111.582 (110.737-112.432) |
| **Wales** |  |  |  |
| 2000-2004 | 921 | 34.324 | 26.832 (25.154-28.623) |
| 2005-2009 | 1690 | 38.271 | 44.159 (42.103-46.316) |
| 2010-2014 | 2581 | 40.647 | 63.498 (61.095-65.996) |
| 2015-2019 | 3914 | 40.116 | 97.567 (94.558-100.672) |
| **Scotland** |  |  |  |
| 2000-2004 | 1388 | 54.525 | 25.456 (24.152-26.831) |
| 2005-2009 | 2729 | 57.039 | 47.845 (46.083-49.674) |
| 2010-2014 | 3935 | 59.420 | 66.223 (64.186-68.325) |
| 2015-2019 | 4617 | 60.950 | 75.750 (73.596-77.967) |
| **Northern Ireland** |  |  |  |
| 2000-2004 | 189 | 12.228 | 15.456 (13.403-17.825) |
| 2005-2009 | 608 | 12.943 | 46.974 (43.385-50.860) |
| 2010-2014 | 951 | 13.500 | 70.444 (66.106-75.066) |
| 2015-2019 | 1455 | 13.782 | 105.569 (100.281-111.135) |

**SA Table 6:** Crude incidence rates by year and country in primary care only study populations (narcolepsy)

| Calendar year | Incident cases (n) | Person years / 100 000 | Crude incidence rate per 100 000 person years (95% CI) |
| --- | --- | --- | --- |
| **United Kingdom** |  |  |  |
| 2000-2004 | 683 | 816.888 | 0.836 (0.776-0.901) |
| 2005-2009 | 727 | 878.156 | 0.828 (0.770-0.890) |
| 2010-2014 | 752 | 900.179 | 0.835 (0.778-0.897) |
| 2015-2019 | 708 | 903.324 | 0.784 (0.728-0.844) |
| **England** |  |  |  |
| 2000-2004 | 584 | 691.087 | 0.845 (0.779-0.916) |
| 2005-2009 | 618 | 743.677 | 0.831 (0.768-0.899) |
| 2010-2014 | 643 | 758.543 | 0.848 (0.785-0.916) |
| 2015-2019 | 624 | 759.485 | 0.822 (0.760-0.889) |
| **Wales** |  |  |  |
| 2000-2004 | 30 | 42.469 | 0.706 (0.494-1.010) |
| 2005-2009 | 38 | 47.292 | 0.804 (0.585-1.104) |
| 2010-2014 | 44 | 50.509 | 0.871 (0.648-1.171) |
| 2015-2019 | 33 | 50.238 | 0.657 (0.467-0.924) |
| **Scotland** |  |  |  |
| 2000-2004 | 54 | 67.222 | 0.803 (0.615-1.049) |
| 2005-2009 | 51 | 70.294 | 0.726 (0.551-0.955) |
| 2010-2014 | 53 | 73.503 | 0.721 (0.551-0.944) |
| 2015-2019 | 46 | 75.548 | 0.609 (0.456-0.813) |
| **Northern Ireland** |  |  |  |
| 2000-2004 | 15 | 16.110 | 0.931 (0.561-1.544) |
| 2005-2009 | 20 | 16.892 | 1.184 (0.764-1.835) |
| 2010-2014 | 12 | 17.624 | 0.681 (0.387-1.199) |
| 2015-2019 | 5 | 18.053 | 0.277 (0.115-0.665) |

**SA Table 7:** Crude and standardised prevalence by year and country in primary care only study populations (obstructive sleep apnoea - OSA)

| Year | Prevalent cases (n) | Study population (n) | Crude prevalence % (95% CI) | Standardised prevalence % (95% CI) | Estimated cases in the population n (95% plausible interval) |
| --- | --- | --- | --- | --- | --- |
| **United Kingdom** |  |  |  |  |  |
| 2000 | 11481 | 12638050 | 0.091 (0.089-0.093) | 0.091 (0.089-0.093) | 41,316 (40,561-42,071) |
| 2001 | 14311 | 12931397 | 0.111 (0.109-0.112) | 0.111 (0.109-0.112) | 50,638 (49,809-51,466) |
| 2002 | 17756 | 13162026 | 0.135 (0.133-0.137) | 0.135 (0.133-0.137) | 62,120 (61,208-63,033) |
| 2003 | 21819 | 13413640 | 0.163 (0.161-0.165) | 0.163 (0.161-0.165) | 75,400 (74,401-76,399) |
| 2004 | 26488 | 13625048 | 0.194 (0.192-0.197) | 0.194 (0.192-0.197) | 90,767 (89,677-91,858) |
| 2005 | 31291 | 13793276 | 0.227 (0.224-0.229) | 0.227 (0.224-0.229) | 106,992 (105,809-108,174) |
| 2006 | 36647 | 13974812 | 0.262 (0.260-0.265) | 0.262 (0.260-0.265) | 124,802 (123,528-126,076) |
| 2007 | 42398 | 14162557 | 0.299 (0.297-0.302) | 0.299 (0.297-0.302) | 143,825 (142,460-145,190) |
| 2008 | 49640 | 14299943 | 0.347 (0.344-0.350) | 0.347 (0.344-0.350) | 168,358 (166,882-169,833) |
| 2009 | 57810 | 14394476 | 0.402 (0.398-0.405) | 0.402 (0.398-0.405) | 196,428 (194,833-198,023) |
| 2010 | 66008 | 14445072 | 0.457 (0.453-0.460) | 0.457 (0.453-0.460) | 225,606 (223,893-227,319) |
| 2011 | 73949 | 14459113 | 0.511 (0.508-0.515) | 0.511 (0.508-0.515) | 254,894 (253,066-256,722) |
| 2012 | 82654 | 14552674 | 0.568 (0.564-0.572) | 0.568 (0.564-0.572) | 285,010 (283,078-286,943) |
| 2013 | 90298 | 14221769 | 0.635 (0.631-0.639) | 0.635 (0.631-0.639) | 320,652 (318,573-322,730) |
| 2014 | 98766 | 14105323 | 0.700 (0.696-0.705) | 0.700 (0.696-0.705) | 356,471 (354,263-358,679) |
| 2015 | 107817 | 14098914 | 0.765 (0.760-0.769) | 0.765 (0.760-0.769) | 392,600 (390,274-394,927) |
| 2016 | 117851 | 14114587 | 0.835 (0.830-0.840) | 0.835 (0.830-0.840) | 432,238 (429,789-434,686) |
| 2017 | 129704 | 14267881 | 0.909 (0.904-0.914) | 0.909 (0.904-0.914) | 473,427 (470,872-475,981) |
| 2018 | 143167 | 14431199 | 0.992 (0.987-0.997) | 0.992 (0.987-0.997) | 519,683 (517,016-522,351) |
| 2019 | 155945 | 14480286 | 1.077 (1.072-1.082) | 1.077 (1.072-1.082) | 567,265 (564,477-570,053) |
| **England** |  |  |  |  |  |
| 2000 | 9662 | 10680146 | 0.090 (0.089-0.093) | 0.091 (0.089-0.092) | 34,447 (33,761-35,133) |
| 2001 | 12068 | 10944400 | 0.110 (0.109-0.112) | 0.110 (0.108-0.112) | 42,282 (41,529-43,036) |
| 2002 | 15071 | 11155898 | 0.135 (0.133-0.137) | 0.135 (0.133-0.137) | 52,176 (51,344-53,008) |
| 2003 | 18575 | 11389635 | 0.163 (0.161-0.165) | 0.163 (0.161-0.166) | 63,450 (62,539-64,361) |
| 2004 | 22518 | 11555450 | 0.195 (0.192-0.197) | 0.195 (0.193-0.198) | 76,372 (75,377-77,368) |
| 2005 | 26583 | 11695614 | 0.227 (0.224-0.229) | 0.228 (0.225-0.231) | 90,056 (88,976-91,136) |
| 2006 | 31154 | 11845333 | 0.263 (0.260-0.265) | 0.264 (0.261-0.267) | 105,190 (104,025-106,355) |
| 2007 | 35925 | 12002535 | 0.299 (0.297-0.302) | 0.300 (0.297-0.304) | 120,870 (119,624-122,116) |
| 2008 | 42198 | 12116052 | 0.348 (0.344-0.350) | 0.350 (0.346-0.353) | 142,061 (140,711-143,412) |
| 2009 | 49208 | 12190743 | 0.404 (0.398-0.405) | 0.406 (0.402-0.409) | 166,142 (164,680-167,604) |
| 2010 | 56116 | 12214594 | 0.459 (0.453-0.460) | 0.462 (0.458-0.466) | 191,002 (189,429-192,575) |
| 2011 | 62833 | 12207407 | 0.515 (0.508-0.515) | 0.518 (0.514-0.522) | 216,154 (214,473-217,836) |
| 2012 | 70296 | 12280638 | 0.572 (0.564-0.572) | 0.576 (0.572-0.580) | 242,219 (240,439-244,000) |
| 2013 | 76524 | 11935135 | 0.641 (0.631-0.639) | 0.645 (0.640-0.650) | 273,234 (271,310-275,158) |
| 2014 | 83598 | 11806362 | 0.708 (0.696-0.705) | 0.712 (0.708-0.717) | 304,346 (302,297-306,396) |
| 2015 | 91362 | 11803821 | 0.774 (0.760-0.769) | 0.779 (0.774-0.784) | 335,657 (333,496-337,818) |
| 2016 | 100054 | 11829250 | 0.846 (0.830-0.840) | 0.851 (0.846-0.857) | 370,166 (367,891-372,442) |
| 2017 | 110341 | 11983845 | 0.921 (0.904-0.914) | 0.927 (0.922-0.933) | 405,694 (403,321-408,068) |
| 2018 | 122087 | 12124239 | 1.007 (0.987-0.997) | 1.015 (1.009-1.020) | 446,657 (444,174-449,139) |
| 2019 | 133036 | 12151605 | 1.095 (1.072-1.082) | 1.104 (1.098-1.110) | 488,662 (486,062-491,262) |
| **Wales** |  |  |  |  |  |
| 2000 | 631 | 651294 | 0.097 (0.089-0.093) | 0.094 (0.087-0.101) | 2,109 (1,945-2,274) |
| 2001 | 766 | 667724 | 0.115 (0.109-0.112) | 0.111 (0.103-0.119) | 2,498 (2,321-2,675) |
| 2002 | 922 | 682012 | 0.135 (0.133-0.137) | 0.131 (0.122-0.139) | 2,963 (2,771-3,154) |
| 2003 | 1149 | 691811 | 0.166 (0.161-0.165) | 0.161 (0.151-0.170) | 3,671 (3,458-3,883) |
| 2004 | 1452 | 715018 | 0.203 (0.192-0.197) | 0.196 (0.186-0.206) | 4,526 (4,292-4,759) |
| 2005 | 1727 | 732836 | 0.236 (0.224-0.229) | 0.227 (0.216-0.237) | 5,263 (5,015-5,512) |
| 2006 | 1990 | 748841 | 0.266 (0.260-0.265) | 0.255 (0.244-0.266) | 5,971 (5,708-6,234) |
| 2007 | 2353 | 763912 | 0.308 (0.297-0.302) | 0.295 (0.283-0.307) | 6,974 (6,692-7,256) |
| 2008 | 2670 | 777488 | 0.343 (0.344-0.350) | 0.329 (0.316-0.341) | 7,849 (7,551-8,147) |
| 2009 | 3041 | 785195 | 0.387 (0.398-0.405) | 0.370 (0.356-0.383) | 8,881 (8,565-9,196) |
| 2010 | 3495 | 799792 | 0.437 (0.453-0.460) | 0.417 (0.403-0.431) | 10,077 (9,743-10,411) |
| 2011 | 3877 | 808406 | 0.480 (0.508-0.515) | 0.457 (0.443-0.471) | 11,110 (10,761-11,460) |
| 2012 | 4311 | 814743 | 0.529 (0.564-0.572) | 0.502 (0.487-0.517) | 12,265 (11,900-12,631) |
| 2013 | 4836 | 818329 | 0.591 (0.631-0.639) | 0.561 (0.545-0.577) | 13,763 (13,376-14,150) |
| 2014 | 5338 | 821261 | 0.650 (0.696-0.705) | 0.617 (0.601-0.634) | 15,201 (14,794-15,607) |
| 2015 | 5683 | 800490 | 0.710 (0.760-0.769) | 0.675 (0.657-0.692) | 16,677 (16,244-17,109) |
| 2016 | 6089 | 788908 | 0.772 (0.830-0.840) | 0.732 (0.714-0.750) | 18,189 (17,734-18,645) |
| 2017 | 6784 | 799511 | 0.849 (0.904-0.914) | 0.802 (0.783-0.821) | 20,024 (19,549-20,499) |
| 2018 | 7554 | 807107 | 0.936 (0.987-0.997) | 0.883 (0.863-0.903) | 22,145 (21,647-22,642) |
| 2019 | 8276 | 812719 | 1.018 (1.072-1.082) | 0.957 (0.937-0.978) | 24,155 (23,637-24,674) |
| **Scotland** |  |  |  |  |  |
| 2000 | 1044 | 1068164 | 0.098 (0.089-0.093) | 0.098 (0.092-0.103) | 3,858 (3,624-4,092) |
| 2001 | 1306 | 1079795 | 0.121 (0.109-0.112) | 0.120 (0.114-0.127) | 4,777 (4,518-5,036) |
| 2002 | 1565 | 1083317 | 0.144 (0.133-0.137) | 0.143 (0.136-0.150) | 5,689 (5,408-5,971) |
| 2003 | 1865 | 1087398 | 0.172 (0.161-0.165) | 0.169 (0.162-0.177) | 6,757 (6,450-7,063) |
| 2004 | 2235 | 1104312 | 0.202 (0.192-0.197) | 0.200 (0.191-0.208) | 8,017 (7,685-8,349) |
| 2005 | 2617 | 1112463 | 0.235 (0.224-0.229) | 0.232 (0.223-0.240) | 9,381 (9,022-9,740) |
| 2006 | 3062 | 1123488 | 0.273 (0.260-0.265) | 0.268 (0.259-0.278) | 10,939 (10,552-11,325) |
| 2007 | 3552 | 1135077 | 0.313 (0.297-0.302) | 0.308 (0.298-0.318) | 12,664 (12,248-13,079) |
| 2008 | 4088 | 1149682 | 0.356 (0.344-0.350) | 0.348 (0.338-0.359) | 14,452 (14,010-14,894) |
| 2009 | 4714 | 1158018 | 0.407 (0.398-0.405) | 0.398 (0.387-0.409) | 16,627 (16,154-17,100) |
| 2010 | 5415 | 1167556 | 0.464 (0.453-0.460) | 0.453 (0.441-0.465) | 19,088 (18,581-19,594) |
| 2011 | 6098 | 1176601 | 0.518 (0.508-0.515) | 0.505 (0.492-0.517) | 21,486 (20,948-22,023) |
| 2012 | 6748 | 1188390 | 0.568 (0.564-0.572) | 0.553 (0.539-0.566) | 23,624 (23,063-24,185) |
| 2013 | 7446 | 1197003 | 0.622 (0.631-0.639) | 0.606 (0.592-0.620) | 26,012 (25,424-26,600) |
| 2014 | 8131 | 1203149 | 0.676 (0.696-0.705) | 0.659 (0.645-0.673) | 28,440 (27,825-29,055) |
| 2015 | 8845 | 1216609 | 0.727 (0.760-0.769) | 0.709 (0.695-0.724) | 30,796 (30,158-31,434) |
| 2016 | 9607 | 1226582 | 0.783 (0.830-0.840) | 0.762 (0.747-0.777) | 33,308 (32,646-33,970) |
| 2017 | 10175 | 1210711 | 0.840 (0.904-0.914) | 0.815 (0.799-0.831) | 35,818 (35,126-36,509) |
| 2018 | 10912 | 1222661 | 0.892 (0.987-0.997) | 0.864 (0.848-0.880) | 38,103 (37,392-38,813) |
| 2019 | 11745 | 1235059 | 0.951 (1.072-1.082) | 0.918 (0.901-0.934) | 40,687 (39,956-41,418) |
| **Northern Ireland** |  |  |  |  |  |
| 2000 | 144 | 238446 | 0.060 (0.089-0.093) | 0.064 (0.053-0.074) | 781 (653-908) |
| 2001 | 171 | 239478 | 0.071 (0.109-0.112) | 0.075 (0.064-0.086) | 925 (786-1,064) |
| 2002 | 198 | 240799 | 0.082 (0.133-0.137) | 0.086 (0.074-0.098) | 1,080 (929-1,230) |
| 2003 | 230 | 244796 | 0.094 (0.161-0.165) | 0.098 (0.085-0.110) | 1,236 (1,076-1,396) |
| 2004 | 283 | 250268 | 0.113 (0.192-0.197) | 0.117 (0.104-0.131) | 1,501 (1,326-1,676) |
| 2005 | 364 | 252363 | 0.144 (0.224-0.229) | 0.149 (0.133-0.164) | 1,926 (1,728-2,124) |
| 2006 | 441 | 257150 | 0.171 (0.260-0.265) | 0.177 (0.160-0.193) | 2,321 (2,104-2,537) |
| 2007 | 568 | 261033 | 0.218 (0.297-0.302) | 0.223 (0.205-0.242) | 2,974 (2,729-3,218) |
| 2008 | 684 | 256721 | 0.266 (0.344-0.350) | 0.272 (0.252-0.293) | 3,672 (3,397-3,947) |
| 2009 | 847 | 260520 | 0.325 (0.398-0.405) | 0.331 (0.308-0.353) | 4,509 (4,206-4,812) |
| 2010 | 982 | 263130 | 0.373 (0.453-0.460) | 0.378 (0.355-0.402) | 5,200 (4,876-5,525) |
| 2011 | 1141 | 266699 | 0.428 (0.508-0.515) | 0.432 (0.407-0.456) | 5,969 (5,624-6,315) |
| 2012 | 1299 | 268903 | 0.483 (0.564-0.572) | 0.484 (0.458-0.510) | 6,735 (6,370-7,100) |
| 2013 | 1492 | 271302 | 0.550 (0.631-0.639) | 0.549 (0.521-0.577) | 7,671 (7,284-8,059) |
| 2014 | 1699 | 274551 | 0.619 (0.696-0.705) | 0.617 (0.588-0.646) | 8,684 (8,273-9,095) |
| 2015 | 1927 | 277994 | 0.693 (0.760-0.769) | 0.690 (0.659-0.720) | 9,776 (9,342-10,210) |
| 2016 | 2101 | 269847 | 0.779 (0.830-0.840) | 0.776 (0.743-0.809) | 11,072 (10,602-11,542) |
| 2017 | 2404 | 273814 | 0.878 (0.904-0.914) | 0.870 (0.836-0.905) | 12,487 (11,991-12,982) |
| 2018 | 2614 | 277192 | 0.943 (0.987-0.997) | 0.931 (0.896-0.967) | 13,443 (12,932-13,954) |
| 2019 | 2888 | 280903 | 1.028 (1.072-1.082) | 1.010 (0.974-1.047) | 14,677 (14,146-15,208) |

**SA Table 8:** Crude and standardised prevalence by year and country in primary care only study populations (narcolepsy)

|  | Prevalent cases (n) | Study population (n) | Crude prevalence % (95% CI) | Standardised prevalence % (95% CI) | Estimated cases in the population n (95% plausible interval) |
| --- | --- | --- | --- | --- | --- |
| **United Kingdom** |  |  |  |  |  |
| 2000 | 1319 | 15690167 | 0.008 (0.008-0.009) | 0.008 (0.008-0.009) | 4,950 (4,683-5,217) |
| 2001 | 1442 | 16055263 | 0.009 (0.009-0.009) | 0.009 (0.009-0.009) | 5,309 (5,035-5,583) |
| 2002 | 1603 | 16333977 | 0.010 (0.009-0.010) | 0.010 (0.009-0.010) | 5,826 (5,541-6,111) |
| 2003 | 1746 | 16648376 | 0.010 (0.010-0.011) | 0.010 (0.010-0.011) | 6,254 (5,961-6,548) |
| 2004 | 1904 | 16916100 | 0.011 (0.011-0.012) | 0.011 (0.011-0.012) | 6,748 (6,445-7,051) |
| 2005 | 2031 | 17136120 | 0.012 (0.011-0.012) | 0.012 (0.011-0.012) | 7,160 (6,849-7,472) |
| 2006 | 2151 | 17363686 | 0.012 (0.012-0.013) | 0.012 (0.012-0.013) | 7,535 (7,217-7,854) |
| 2007 | 2288 | 17606086 | 0.013 (0.012-0.014) | 0.013 (0.012-0.014) | 7,969 (7,642-8,295) |
| 2008 | 2431 | 17796617 | 0.014 (0.013-0.014) | 0.014 (0.013-0.014) | 8,445 (8,109-8,781) |
| 2009 | 2500 | 17941081 | 0.014 (0.013-0.014) | 0.014 (0.013-0.014) | 8,676 (8,336-9,016) |
| 2010 | 2613 | 18019128 | 0.015 (0.014-0.015) | 0.015 (0.014-0.015) | 9,101 (8,752-9,450) |
| 2011 | 2674 | 18079655 | 0.015 (0.014-0.015) | 0.015 (0.014-0.015) | 9,360 (9,005-9,715) |
| 2012 | 2778 | 18230035 | 0.015 (0.015-0.016) | 0.015 (0.015-0.016) | 9,708 (9,347-10,069) |
| 2013 | 2802 | 17889030 | 0.016 (0.015-0.016) | 0.016 (0.015-0.016) | 10,041 (9,669-10,413) |
| 2014 | 2883 | 17797535 | 0.016 (0.016-0.017) | 0.016 (0.016-0.017) | 10,464 (10,082-10,846) |
| 2015 | 2909 | 17813561 | 0.016 (0.016-0.017) | 0.016 (0.016-0.017) | 10,633 (10,246-11,019) |
| 2016 | 2942 | 17860814 | 0.016 (0.016-0.017) | 0.016 (0.016-0.017) | 10,813 (10,423-11,204) |
| 2017 | 2990 | 18075482 | 0.017 (0.016-0.017) | 0.017 (0.016-0.017) | 10,924 (10,533-11,316) |
| 2018 | 3102 | 18284633 | 0.017 (0.016-0.018) | 0.017 (0.016-0.018) | 11,271 (10,874-11,667) |
| 2019 | 3144 | 18342324 | 0.017 (0.017-0.018) | 0.017 (0.017-0.018) | 11,449 (11,049-11,850) |
| **England** |  |  |  |  |  |
| 2000 | 1119 | 13237155 | 0.008 (0.008-0.009) | 0.008 (0.008-0.009) | 4,158 (3,914-4,402) |
| 2001 | 1219 | 13566851 | 0.009 (0.009-0.009) | 0.009 (0.008-0.009) | 4,441 (4,192-4,690) |
| 2002 | 1362 | 13823413 | 0.010 (0.009-0.010) | 0.010 (0.009-0.010) | 4,894 (4,634-5,153) |
| 2003 | 1487 | 14116626 | 0.011 (0.010-0.011) | 0.011 (0.010-0.011) | 5,260 (4,993-5,527) |
| 2004 | 1621 | 14329760 | 0.011 (0.011-0.012) | 0.011 (0.011-0.012) | 5,680 (5,404-5,957) |
| 2005 | 1717 | 14517222 | 0.012 (0.011-0.012) | 0.012 (0.011-0.012) | 5,991 (5,708-6,274) |
| 2006 | 1817 | 14707625 | 0.012 (0.012-0.013) | 0.012 (0.012-0.013) | 6,303 (6,013-6,592) |
| 2007 | 1925 | 14914190 | 0.013 (0.012-0.014) | 0.013 (0.012-0.013) | 6,640 (6,343-6,936) |
| 2008 | 2052 | 15073902 | 0.014 (0.013-0.014) | 0.014 (0.013-0.014) | 7,063 (6,757-7,369) |
| 2009 | 2105 | 15192012 | 0.014 (0.013-0.014) | 0.014 (0.013-0.014) | 7,243 (6,933-7,552) |
| 2010 | 2205 | 15237378 | 0.014 (0.014-0.015) | 0.014 (0.014-0.015) | 7,631 (7,312-7,949) |
| 2011 | 2252 | 15269404 | 0.015 (0.014-0.015) | 0.015 (0.014-0.015) | 7,845 (7,521-8,169) |
| 2012 | 2337 | 15393035 | 0.015 (0.015-0.016) | 0.015 (0.015-0.016) | 8,136 (7,807-8,466) |
| 2013 | 2348 | 15031848 | 0.016 (0.015-0.016) | 0.016 (0.015-0.016) | 8,434 (8,093-8,775) |
| 2014 | 2406 | 14923577 | 0.016 (0.016-0.017) | 0.016 (0.016-0.017) | 8,782 (8,431-9,132) |
| 2015 | 2439 | 14943195 | 0.016 (0.016-0.017) | 0.016 (0.016-0.017) | 8,968 (8,612-9,324) |
| 2016 | 2474 | 15002301 | 0.016 (0.016-0.017) | 0.017 (0.016-0.017) | 9,144 (8,784-9,504) |
| 2017 | 2520 | 15219470 | 0.017 (0.016-0.017) | 0.017 (0.016-0.017) | 9,241 (8,880-9,602) |
| 2018 | 2613 | 15402023 | 0.017 (0.016-0.018) | 0.017 (0.016-0.018) | 9,530 (9,164-9,895) |
| 2019 | 2654 | 15434759 | 0.017 (0.017-0.018) | 0.017 (0.017-0.018) | 9,713 (9,343-10,082) |
| **Wales** |  |  |  |  |  |
| 2000 | 61 | 811504 | 0.008 (0.008-0.009) | 0.007 (0.005-0.009) | 212 (159-265) |
| 2001 | 67 | 831795 | 0.008 (0.009-0.009) | 0.008 (0.006-0.010) | 226 (172-281) |
| 2002 | 72 | 848153 | 0.008 (0.009-0.010) | 0.008 (0.006-0.010) | 240 (184-295) |
| 2003 | 76 | 859683 | 0.009 (0.010-0.011) | 0.009 (0.007-0.010) | 251 (195-308) |
| 2004 | 84 | 887228 | 0.009 (0.011-0.012) | 0.009 (0.007-0.011) | 271 (213-329) |
| 2005 | 93 | 908639 | 0.010 (0.011-0.012) | 0.010 (0.008-0.012) | 293 (233-352) |
| 2006 | 102 | 928687 | 0.011 (0.012-0.013) | 0.011 (0.009-0.013) | 316 (255-378) |
| 2007 | 111 | 947076 | 0.012 (0.012-0.014) | 0.011 (0.009-0.013) | 341 (277-405) |
| 2008 | 113 | 964451 | 0.012 (0.013-0.014) | 0.011 (0.009-0.014) | 345 (281-409) |
| 2009 | 118 | 974705 | 0.012 (0.013-0.014) | 0.012 (0.010-0.014) | 358 (293-423) |
| 2010 | 120 | 992503 | 0.012 (0.014-0.015) | 0.012 (0.010-0.014) | 357 (293-421) |
| 2011 | 126 | 1004425 | 0.013 (0.014-0.015) | 0.012 (0.010-0.014) | 372 (307-437) |
| 2012 | 135 | 1013274 | 0.013 (0.015-0.016) | 0.013 (0.011-0.015) | 397 (330-464) |
| 2013 | 137 | 1019251 | 0.013 (0.015-0.016) | 0.013 (0.011-0.015) | 400 (333-467) |
| 2014 | 154 | 1024517 | 0.015 (0.016-0.017) | 0.014 (0.012-0.017) | 447 (376-518) |
| 2015 | 151 | 998908 | 0.015 (0.016-0.017) | 0.015 (0.012-0.017) | 451 (379-524) |
| 2016 | 149 | 985176 | 0.015 (0.016-0.017) | 0.015 (0.012-0.017) | 454 (381-528) |
| 2017 | 151 | 999326 | 0.015 (0.016-0.017) | 0.015 (0.012-0.017) | 454 (382-527) |
| 2018 | 161 | 1009351 | 0.016 (0.016-0.018) | 0.015 (0.013-0.018) | 484 (408-559) |
| 2019 | 159 | 1015962 | 0.016 (0.017-0.018) | 0.015 (0.013-0.018) | 479 (404-554) |
| **Scotland** |  |  |  |  |  |
| 2000 | 120 | 1324178 | 0.009 (0.008-0.009) | 0.009 (0.007-0.011) | 462 (379-545) |
| 2001 | 135 | 1337916 | 0.010 (0.009-0.009) | 0.010 (0.008-0.012) | 515 (428-602) |
| 2002 | 147 | 1342477 | 0.011 (0.009-0.010) | 0.011 (0.009-0.013) | 559 (468-649) |
| 2003 | 158 | 1347412 | 0.012 (0.010-0.011) | 0.012 (0.010-0.014) | 597 (503-690) |
| 2004 | 171 | 1367629 | 0.013 (0.011-0.012) | 0.013 (0.011-0.014) | 640 (544-737) |
| 2005 | 189 | 1376731 | 0.014 (0.011-0.012) | 0.014 (0.012-0.016) | 708 (607-809) |
| 2006 | 196 | 1389048 | 0.014 (0.012-0.013) | 0.014 (0.012-0.016) | 727 (625-828) |
| 2007 | 210 | 1402461 | 0.015 (0.012-0.014) | 0.015 (0.013-0.017) | 775 (670-880) |
| 2008 | 221 | 1421974 | 0.016 (0.013-0.014) | 0.016 (0.013-0.018) | 808 (701-915) |
| 2009 | 230 | 1433083 | 0.016 (0.013-0.014) | 0.016 (0.014-0.018) | 837 (729-946) |
| 2010 | 238 | 1444595 | 0.016 (0.014-0.015) | 0.016 (0.014-0.018) | 862 (752-971) |
| 2011 | 245 | 1456576 | 0.017 (0.014-0.015) | 0.017 (0.015-0.019) | 884 (773-995) |
| 2012 | 251 | 1471450 | 0.017 (0.015-0.016) | 0.017 (0.015-0.019) | 900 (789-1,012) |
| 2013 | 260 | 1482537 | 0.018 (0.015-0.016) | 0.017 (0.015-0.019) | 926 (813-1,039) |
| 2014 | 268 | 1489930 | 0.018 (0.016-0.017) | 0.018 (0.016-0.020) | 953 (839-1,067) |
| 2015 | 266 | 1507362 | 0.018 (0.016-0.017) | 0.017 (0.015-0.020) | 938 (825-1,051) |
| 2016 | 270 | 1519442 | 0.018 (0.016-0.017) | 0.018 (0.015-0.020) | 946 (833-1,059) |
| 2017 | 269 | 1497834 | 0.018 (0.016-0.017) | 0.018 (0.015-0.020) | 955 (840-1,069) |
| 2018 | 278 | 1510716 | 0.018 (0.016-0.018) | 0.018 (0.016-0.020) | 979 (863-1,094) |
| 2019 | 283 | 1524757 | 0.019 (0.017-0.018) | 0.018 (0.016-0.020) | 990 (874-1,105) |
| **Northern Ireland** |  |  |  |  |  |
| 2000 | 19 | 317330 | 0.006 (0.008-0.009) | 0.007 (0.004-0.010) | 116 (64-169) |
| 2001 | 21 | 318701 | 0.007 (0.009-0.009) | 0.007 (0.004-0.011) | 126 (72-180) |
| 2002 | 22 | 319934 | 0.007 (0.009-0.010) | 0.008 (0.004-0.011) | 132 (76-187) |
| 2003 | 25 | 324655 | 0.008 (0.010-0.011) | 0.008 (0.005-0.012) | 145 (88-201) |
| 2004 | 28 | 331483 | 0.008 (0.011-0.012) | 0.009 (0.006-0.013) | 158 (99-217) |
| 2005 | 32 | 333528 | 0.010 (0.011-0.012) | 0.010 (0.007-0.014) | 181 (118-244) |
| 2006 | 36 | 338326 | 0.011 (0.012-0.013) | 0.012 (0.008-0.015) | 202 (136-268) |
| 2007 | 42 | 342359 | 0.012 (0.012-0.014) | 0.013 (0.009-0.017) | 235 (163-306) |
| 2008 | 45 | 336290 | 0.013 (0.013-0.014) | 0.014 (0.010-0.019) | 255 (180-330) |
| 2009 | 47 | 341281 | 0.014 (0.013-0.014) | 0.015 (0.010-0.019) | 264 (188-339) |
| 2010 | 50 | 344652 | 0.015 (0.014-0.015) | 0.015 (0.011-0.020) | 279 (201-356) |
| 2011 | 51 | 349250 | 0.015 (0.014-0.015) | 0.015 (0.011-0.020) | 280 (203-357) |
| 2012 | 55 | 352276 | 0.016 (0.015-0.016) | 0.016 (0.012-0.021) | 299 (220-378) |
| 2013 | 57 | 355394 | 0.016 (0.015-0.016) | 0.017 (0.012-0.021) | 305 (226-385) |
| 2014 | 55 | 359511 | 0.015 (0.016-0.017) | 0.016 (0.012-0.020) | 292 (215-369) |
| 2015 | 53 | 364096 | 0.015 (0.016-0.017) | 0.015 (0.011-0.019) | 277 (202-352) |
| 2016 | 49 | 353895 | 0.014 (0.016-0.017) | 0.014 (0.010-0.018) | 265 (191-339) |
| 2017 | 50 | 358852 | 0.014 (0.016-0.017) | 0.014 (0.010-0.018) | 267 (193-341) |
| 2018 | 50 | 362543 | 0.014 (0.016-0.018) | 0.014 (0.010-0.018) | 267 (193-341) |
| 2019 | 48 | 366846 | 0.013 (0.017-0.018) | 0.013 (0.010-0.017) | 254 (182-326) |

**SA Table 6:**
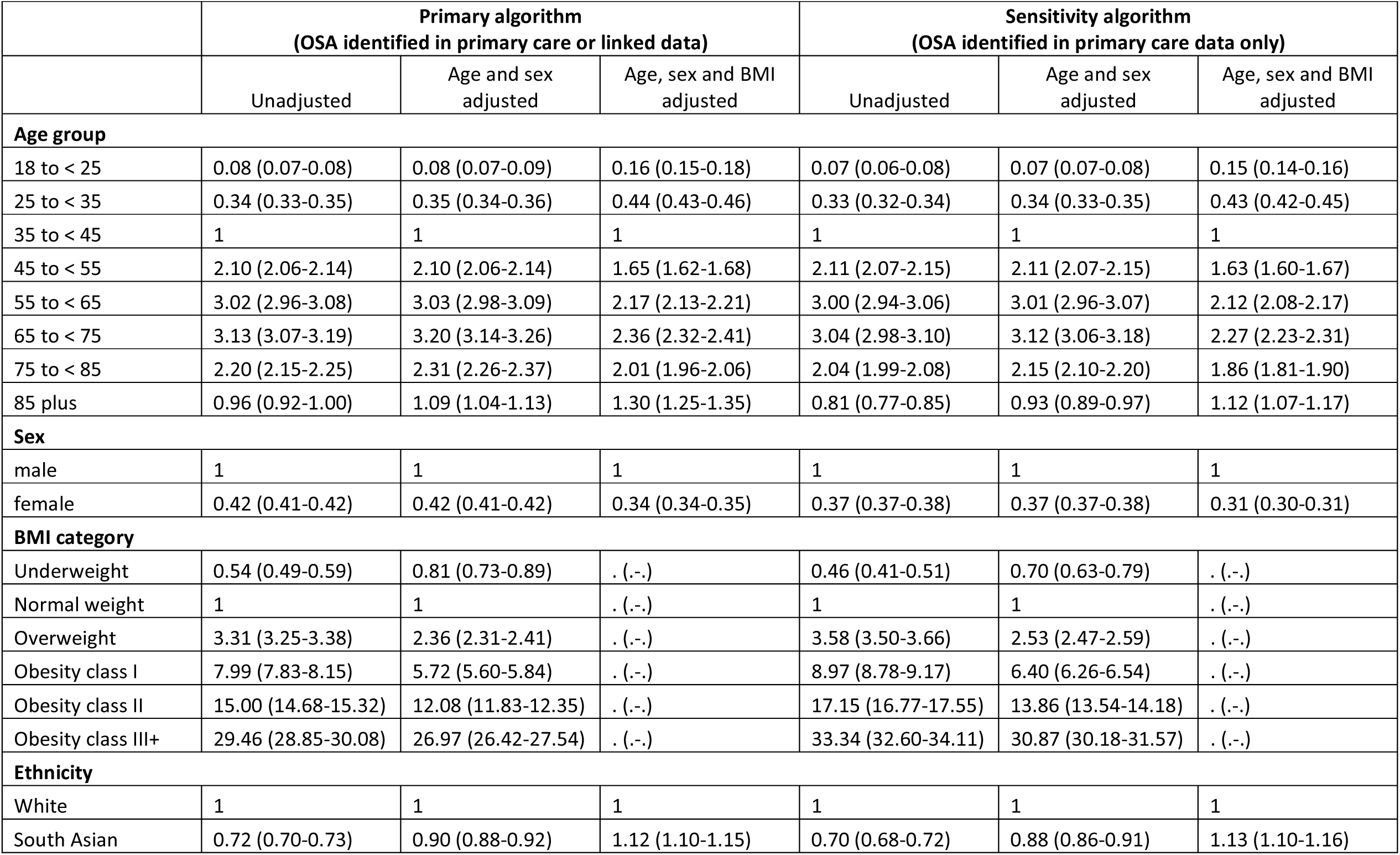
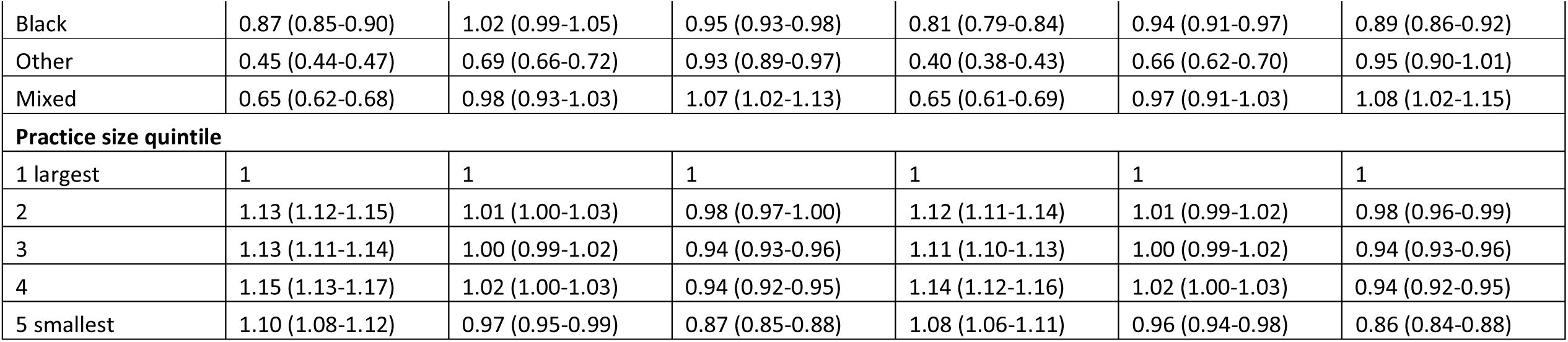
2019 Prevalence Ratios in England using the primary linked algorithm and primary care only algorithms as a sensitivity analysis (OSA) Caption (SA tables 6-9): Deprivation and urban rural data were not available for the full primary care population.

**SA Table 7:** 2019 Incidence Rate Ratios in England using the primary linked algorithm and primary care only algorithms as a sensitivity analysis (OSA)

|  | Primary algorithm<br>(OSA identified in primary care or linked data) |  |  | Sensitivity algorithm<br>(OSA identified in primary care data only) |  |  |
| --- | --- | --- | --- | --- | --- | --- |
|  | Unadjusted | Age and sex<br>adjusted | Age, sex and BMI<br>adjusted | Unadjusted | Age and sex<br>adjusted | Age, sex and BMI<br>adjusted |
| **Age group** |  |  |  |  |  |  |
| 18 to < 25 | 0.20 (0.18-0.22) | 0.20 (0.18-0.23) | 0.36 (0.32-0.42) | 0.11 (0.10-0.12) | 0.11 (0.10-0.13) | 0.20 (0.17-0.23) |
| 25 to < 35 | 0.50 (0.47-0.53) | 0.50 (0.47-0.53) | 0.60 (0.56-0.64) | 0.34 (0.31-0.36) | 0.35 (0.33-0.37) | 0.44 (0.41-0.47) |
| 35 to < 45 | 1 | 1 | 1 | 1 | 1 | 1 |
| 45 to < 55 | 1.56 (1.49-1.64) | 1.57 (1.49-1.64) | 1.26 (1.20-1.32) | 1.93 (1.84-2.03) | 1.91 (1.82-2.00) | 1.48 (1.41-1.55) |
| 55 to < 65 | 1.88 (1.79-1.97) | 1.89 (1.80-1.98) | 1.41 (1.34-1.48) | 2.26 (2.15-2.37) | 2.25 (2.14-2.36) | 1.61 (1.53-1.69) |
| 65 to < 75 | 1.64 (1.56-1.73) | 1.67 (1.58-1.76) | 1.29 (1.22-1.36) | 1.67 (1.58-1.76) | 1.66 (1.58-1.76) | 1.22 (1.16-1.30) |
| 75 to < 85 | 1.17 (1.10-1.25) | 1.22 (1.14-1.30) | 1.09 (1.02-1.16) | 0.75 (0.70-0.81) | 0.76 (0.71-0.82) | 0.68 (0.63-0.73) |
| 85 plus | 0.49 (0.43-0.56) | 0.53 (0.47-0.61) | 0.65 (0.56-0.74) | 0.12 (0.10-0.14) | 0.13 (0.11-0.16) | 0.18 (0.15-0.21) |
| **Sex** |  |  |  |  |  |  |
| male | 1 | 1 | 1 | 1 | 1 | 1 |
| female | 0.55 (0.54-0.57) | 0.56 (0.54-0.57) | 0.43 (0.42-0.45) | 0.45 (0.44-0.47) | 0.52 (0.50-0.54) | 0.44 (0.43-0.46) |
| **BMI category** |  |  |  |  |  |  |
| Normal weight | 1 | 1 | . (-.) | 1 | 1 | . (-.) |
| Overweight | 2.80 (2.64-2.96) | 2.24 (2.11-2.37) | . (-.) | 3.69 (3.48-3.92) | 2.62 (2.46-2.78) | . (-.) |
| Obesity class I | 6.60 (6.24-6.99) | 5.39 (5.09-5.70) | . (-.) | 9.27 (8.73-9.84) | 6.38 (6.01-6.78) | . (-.) |
| Obesity class II | 13.61 (12.83-14.44) | 12.23 (11.52-12.98) | . (-.) | 18.01 (16.91-19.18) | 13.05 (12.25-13.91) | . (-.) |
| Obesity class III+ | 30.10 (28.41-31.89) | 30.00 (28.30-31.80) | . (-.) | 36.13 (33.97-38.42) | 28.01 (26.32-29.81) | . (-.) |
| **Ethnicity** |  |  |  |  |  |  |
| White | 1 | 1 | 1 | 1 | 1 | 1 |
| South Asian | 0.85 (0.80-0.90) | 0.95 (0.89-1.01) | 1.16 (1.09-1.23) | 1.02 (0.96-1.10) | 1.16 (1.08-1.24) | 1.42 (1.32-1.52) |
| Black | 0.89 (0.82-0.96) | 0.94 (0.87-1.02) | 0.86 (0.79-0.92) | 0.93 (0.85-1.02) | 0.97 (0.88-1.06) | 0.91 (0.83-1.00) |
| Other | 0.54 (0.48-0.60) | 0.68 (0.61-0.76) | 0.92 (0.82-1.03) | 0.35 (0.30-0.41) | 0.57 (0.49-0.67) | 0.82 (0.70-0.95) |
| Mixed | 0.71 (0.62-0.82) | 0.91 (0.79-1.05) | 0.99 (0.85-1.15) | 0.56 (0.47-0.67) | 0.77 (0.64-0.92) | 0.87 (0.73-1.04) |
| **Practice size quintile** |  |  |  |  |  |  |
| 1 largest | 1 | 1 | 1 | 1 | 1 | 1 |
| 2 | 1.11 (1.07-1.16) | 1.03 (0.99-1.07) | 0.99 (0.95-1.03) | 1.36 (1.31-1.42) | 1.16 (1.12-1.21) | 1.11 (1.06-1.16) |
| 3 | 1.10 (1.06-1.15) | 1.02 (0.98-1.07) | 0.95 (0.91-0.99) | 1.13 (1.08-1.18) | 0.92 (0.88-0.96) | 0.88 (0.84-0.92) |
| 4 | 1.17 (1.11-1.22) | 1.08 (1.03-1.13) | 0.98 (0.93-1.03) | 1.34 (1.27-1.40) | 1.07 (1.02-1.12) | 0.97 (0.92-1.02) |
| 5 smallest | 1.06 (1.00-1.12) | 0.97 (0.91-1.03) | 0.84 (0.79-0.90) | 1.22 (1.15-1.30) | 0.98 (0.92-1.04) | 0.84 (0.79-0.90) |

**SA Table 8:** 2015-2019 Prevalence Ratios in in England using the primary linked algorithm and primary care only algorithms as a sensitivity analysis (narcolepsy)

|  | **Primary algorithm**<br>**(narcolepsy identified in primary**<br>**care or linked data)** |  | **Sensitivity algorithm**<br>**(narcolepsy identified in primary**<br>**care data only)** |  |
| --- | --- | --- | --- | --- |
|  | Unadjusted | Age and sex<br>adjusted | Unadjusted | Age and sex<br>adjusted |
| **Age group** |  |  |  |  |
| 0 to < 9 | 0.06 (0.04-0.09) | 0.06 (0.04-0.09) | 0.05 (0.03-0.07) | 0.05 (0.03-0.07) |
| 9 to < 18 | 0.28 (0.23-0.34) | 0.28 (0.23-0.34) | 0.28 (0.22-0.34) | 0.28 (0.22-0.34) |
| 18 to < 25 | 0.52 (0.44-0.62) | 0.52 (0.43-0.62) | 0.51 (0.43-0.61) | 0.51 (0.43-0.61) |
| 25 to < 35 | 0.74 (0.65-0.84) | 0.74 (0.65-0.84) | 0.73 (0.64-0.84) | 0.73 (0.64-0.83) |
| 35 to < 45 | 1 | 1 | 1 | 1 |
| 45 to < 55 | 0.97 (0.86-1.10) | 0.97 (0.86-1.10) | 0.95 (0.84-1.08) | 0.95 (0.84-1.08) |
| 55 to < 65 | 0.84 (0.74-0.96) | 0.84 (0.73-0.96) | 0.86 (0.75-0.98) | 0.86 (0.75-0.98) |
| 65 to < 75 | 0.84 (0.72-0.97) | 0.84 (0.72-0.97) | 0.86 (0.75-1.00) | 0.86 (0.74-0.99) |
| 75 to < 85 | 1.07 (0.92-1.26) | 1.06 (0.91-1.25) | 1.16 (0.99-1.35) | 1.15 (0.99-1.34) |
| 85 plus | 1.23 (1.00-1.51) | 1.21 (0.98-1.49) | 1.15 (0.93-1.42) | 1.14 (0.92-1.41) |
| **Sex** |  |  |  |  |
| male | 1 | 1 | 1 | 1 |
| female | 1.15 (1.06-1.24) | 1.13 (1.05-1.22) | 1.08 (1.00-1.16) | 1.06 (0.98-1.15) |
| **Ethnicity** |  |  |  |  |
| White | 1 | 1 | 1 | 1 |
| South Asian | 0.33 (0.27-0.42) | 0.36 (0.29-0.46) | 0.40 (0.32-0.51) | 0.43 (0.34-0.55) |
| Black | 1.40 (1.20-1.62) | 1.51 (1.30-1.75) | 1.29 (1.08-1.55) | 1.39 (1.16-1.67) |
| Other | 0.31 (0.22-0.46) | 0.34 (0.23-0.49) | 0.30 (0.19-0.48) | 0.32 (0.20-0.52) |
| Mixed | 0.91 (0.69-1.20) | 1.24 (0.94-1.63) | 0.87 (0.62-1.22) | 1.12 (0.80-1.57) |
| **Practice size quintile** |  |  |  |  |
| 1 largest | 1 | 1 | 1 | 1 |
| 2 | 1.09 (0.99-1.20) | 1.07 (0.97-1.18) | 1.05 (0.95-1.16) | 1.03 (0.93-1.14) |
| 3 | 1.01 (0.90-1.12) | 0.99 (0.88-1.10) | 0.95 (0.85-1.05) | 0.93 (0.83-1.03) |
| 4 | 0.96 (0.85-1.09) | 0.95 (0.84-1.08) | 0.87 (0.77-0.99) | 0.86 (0.76-0.98) |
| 5 smallest | 0.95 (0.82-1.11) | 0.95 (0.81-1.11) | 0.94 (0.81-1.09) | 0.93 (0.80-1.08) |

**SA Table 9:** 2015-2019 Incidence Rate Ratios in England using the primary linked algorithm and primary care only algorithms as a sensitivity analysis (narcolepsy)

|  | Primary algorithm<br>(narcolepsy identified in primary<br>care or linked data) |  | Sensitivity algorithm<br>(narcolepsy identified in primary<br>care data only) |  |
| --- | --- | --- | --- | --- |
|  | Unadjusted | Age and sex<br>adjusted | Unadjusted | Age and sex<br>adjusted |
| **Age group** |  |  |  |  |
| 0 to < 9 | 0.31 (0.21-0.46) | 0.31 (0.21-0.46) | 0.35 (0.23-0.52) | 0.35 (0.23-0.52) |
| 9 to < 18 | 0.90 (0.69-1.17) | 0.90 (0.69-1.17) | 0.97 (0.73-1.29) | 0.97 (0.73-1.29) |
| 18 to < 25 | 1.12 (0.86-1.46) | 1.12 (0.86-1.46) | 1.40 (1.07-1.83) | 1.39 (1.07-1.82) |
| 25 to < 35 | 1.04 (0.82-1.31) | 1.04 (0.82-1.31) | 1.04 (0.81-1.34) | 1.04 (0.81-1.34) |
| 35 to < 45 | 1 | 1 | 1 | 1 |
| 45 to < 55 | 0.78 (0.60-1.00) | 0.78 (0.60-1.00) | 0.63 (0.47-0.84) | 0.62 (0.47-0.84) |
| 55 to < 65 | 0.62 (0.47-0.83) | 0.62 (0.47-0.83) | 0.50 (0.36-0.70) | 0.50 (0.36-0.70) |
| 65 to < 75 | 0.49 (0.34-0.69) | 0.48 (0.34-0.68) | 0.35 (0.23-0.54) | 0.35 (0.23-0.53) |
| 75 to < 85 | 0.62 (0.42-0.91) | 0.61 (0.42-0.90) | 0.28 (0.16-0.50) | 0.28 (0.16-0.49) |
| 85 plus | 0.82 (0.50-1.32) | 0.79 (0.49-1.27) | 0.49 (0.26-0.94) | 0.47 (0.25-0.90) |
| **Sex** |  |  |  |  |
| male | 1 | 1 | 1 | 1 |
| female | 1.27 (1.10-1.46) | 1.28 (1.11-1.47) | 1.33 (1.14-1.56) | 1.35 (1.15-1.59) |
| **Ethnicity** |  |  |  |  |
| White | 1 | 1 | 1 | 1 |
| South Asian | 0.52 (0.36-0.74) | 0.49 (0.34-0.70) | 0.56 (0.36-0.87) | 0.50 (0.32-0.78) |
| Black | 1.75 (1.35-2.28) | 1.67 (1.28-2.17) | 1.96 (1.42-2.71) | 1.77 (1.28-2.45) |
| Other | 0.64 (0.38-1.09) | 0.57 (0.34-0.97) | 0.70 (0.35-1.41) | 0.56 (0.28-1.13) |
| Mixed | 1.44 (0.93-2.22) | 1.44 (0.93-2.23) | 1.99 (1.21-3.29) | 1.79 (1.08-2.96) |
| **Practice size quintile** |  |  |  |  |
| 1 largest | 1 | 1 | 1 | 1 |
| 2 | 1.05 (0.88-1.27) | 1.09 (0.91-1.31) | 0.83 (0.67-1.02) | 0.88 (0.71-1.08) |
| 3 | 0.95 (0.77-1.17) | 0.99 (0.80-1.21) | 0.79 (0.63-0.99) | 0.84 (0.67-1.05) |
| 4 | 1.06 (0.85-1.32) | 1.10 (0.88-1.37) | 0.80 (0.62-1.03) | 0.85 (0.66-1.09) |
| 5 smallest | 0.92 (0.69-1.23) | 0.96 (0.72-1.28) | 0.87 (0.64-1.18) | 0.92 (0.68-1.25) |

**SA Table 10:** Age-sex adjusted prevalence ratios and incidence rate ratios of OSA in the full study population and restricted to people with complete case BMI data.

| Characteristic | Age-sex adjusted Prevalence Ratio |  | Age-sex adjusted Incidence Rate Ratio |  |
| --- | --- | --- | --- | --- |
|  | full study population | complete case | full study population | complete case |
| **Age group** |  |  |  |  |
| 18 to < 25 | 0.08 (0.07-0.09) | 0.12 (0.11-0.13) | 0.20 (0.18-0.23) | 0.27 (0.24-0.31) |
| 25 to < 35 | 0.35 (0.34-0.36) | 0.38 (0.37-0.39) | 0.50 (0.47-0.53) | 0.53 (0.50-0.57) |
| 35 to < 45 | 1 | 1 | 1 | 1 |
| 45 to < 55 | 2.10 (2.06-2.14) | 1.99 (1.96-2.03) | 1.57 (1.49-1.64) | 1.49 (1.42-1.57) |
| 55 to < 65 | 3.03 (2.98-3.09) | 2.83 (2.77-2.88) | 1.89 (1.80-1.98) | 1.77 (1.68-1.86) |
| 65 to < 75 | 3.20 (3.14-3.26) | 2.91 (2.86-2.97) | 1.67 (1.58-1.76) | 1.53 (1.45-1.61) |
| 75 to < 85 | 2.31 (2.26-2.37) | 2.08 (2.04-2.13) | 1.22 (1.14-1.30) | 1.11 (1.03-1.18) |
| 85 plus | 1.09 (1.04-1.13) | 1.00 (0.96-1.04) | 0.53 (0.47-0.61) | 0.50 (0.44-0.57) |
| **Sex** |  |  |  |  |
| male | 1 | 1 | 1 | 1 |
| female | 0.42 (0.41-0.42) | 0.40 (0.39-0.40) | 0.56 (0.54-0.57) | 0.53 (0.52-0.55) |
| **Ethnicity** |  |  |  |  |
| 0. White | 1 | 1 | 1 | 1 |
| 1. South Asian | 0.90 (0.88-0.92) | 0.89 (0.87-0.91) | 0.95 (0.89-1.01) | 0.93 (0.87-0.99) |
| 2. Black | 1.02 (0.99-1.05) | 1.00 (0.97-1.03) | 0.94 (0.87-1.02) | 0.93 (0.86-1.00) |
| 3. Other | 0.69 (0.66-0.72) | 0.69 (0.66-0.72) | 0.68 (0.61-0.76) | 0.68 (0.61-0.77) |
| 4. Mixed | 0.98 (0.93-1.03) | 0.97 (0.92-1.02) | 0.91 (0.79-1.05) | 0.91 (0.78-1.05) |
| **Area based deprivation** |  |  |  |  |
| 1 (lowest) | 1 | 1 | 1 | 1 |
| 2 | 0.98 (0.97-1.00) | 0.98 (0.97-1.00) | 1.02 (0.97-1.08) | 1.03 (0.98-1.09) |
| 3 | 1.04 (1.02-1.06) | 1.04 (1.02-1.06) | 1.06 (1.01-1.12) | 1.07 (1.02-1.13) |
| 4 | 1.13 (1.11-1.15) | 1.13 (1.11-1.15) | 1.19 (1.13-1.25) | 1.20 (1.14-1.27) |
| 5 (highest) | 1.11 (1.09-1.13) | 1.10 (1.08-1.12) | 1.16 (1.11-1.23) | 1.18 (1.12-1.25) |
| **Urban-rural** |  |  |  |  |
| urban | 1 | 1 | 1 | 1 |
| rural | 0.93 (0.91-0.94) | 0.93 (0.92-0.95) | 0.87 (0.83-0.90) | 0.87 (0.83-0.90) |
| **Practice size quintile** |  |  |  |  |
| 1 largest | 1 | 1 | 1 | 1 |
| 2 | 1.01 (1.00-1.03) | 1.01 (1.00-1.03) | 1.03 (0.99-1.07) | 1.03 (0.99-1.08) |
| 3 | 1.00 (0.99-1.02) | 1.00 (0.99-1.02) | 1.02 (0.98-1.07) | 1.02 (0.98-1.07) |
| 4 | 1.02 (1.00-1.03) | 1.01 (0.99-1.03) | 1.08 (1.03-1.13) | 1.08 (1.03-1.13) |
| 5 smallest | 0.97 (0.95-0.99) | 0.96 (0.94-0.98) | 0.97 (0.91-1.03) | 0.95 (0.90-1.01) |

